# Integrating Genetic, Environmental, Cognitive, and Temperament Data for ADHD Prediction in Explainable Deep Learning Models

**DOI:** 10.64898/2026.06.29.26356796

**Authors:** Eric J. Barnett, Michael A. Mooney, Yanli Zhang James, Peter Ryabinin, Stephen V. Faraone

## Abstract

**Objective:** Attention-deficit/hyperactivity disorder (ADHD) is clinically and etiologically heterogeneous, and diagnostic decisions may benefit from integrating multiple sources of information. We developed an explainable deep learning approach to test whether genetic, environmental, cognitive, demographic, and temperament data could classify ADHD diagnosis and identify features contributing to model decisions.

**Method:** We analyzed participants from the Oregon ADHD-1000 cohort split into training, validation, and test subsets. We trained modular neural network models classifying ADHD case-control status using SNP-level genotype data with biological annotations, polygenic scores, demographics, parenting and family conflict, stress and trauma, geocoded measures, cognitive task measures, temperament factor scores, and missingness indicators. Hyperparameter optimization selected model architecture and feature block inclusion. We evaluated model performance using AUC, precision-recall curves, calibration analyses, prediction certainty analyses, and decision curve analysis. We used integrated gradients to quantify block-level, feature-level, and individualized feature importance.

**Results:** The best model using temperament features had an AUC of 0.97 in the held-out test subset, with high accuracy, sensitivity, and specificity and a Brier score of 0.06. The best model excluding temperament had an AUC of 0.75. Feature importance analyses highlighted temperament, demographic, and cognitive domains in the temperament-inclusive model.

Individualized explanations showed that prediction drivers varied across participants and could help reveal conflicting or supporting evidence across domains.

**Conclusion:** Explainable, multi-modal classification models can integrate heterogeneous ADHD-relevant information and identify features that contribute to individual predictions. These types of models may advance ADHD risk modeling research and clinician-led decision support, especially in complex or diagnostically uncertain cases.

## Introduction

Attention-deficit/hyperactivity disorder (ADHD) is a highly heritable and polygenic neurodevelopmental disorder characterized by substantial clinical and etiologic heterogeneity.^1^ Genome-wide association studies (GWAS) have identified common variant associations contributing to ADHD liability, and polygenic risk scores (PRSs) capture a measurable portion of aggregate genetic risk. ^2^ However, predictive performance based solely on common variant information remains modest. Although SNP-based heritability estimates and twin-studies indicate substantial genetic contribution^3^, the translation of genetic risk patterns into accurate individual-level prediction has proven challenging.

In parallel with advances in genomic discovery, environmental and developmental factors have been consistently associated with ADHD risk although causality has been difficult to establish due to genetic confounding.^4^ Parenting practices, family conflict, socioeconomic and geospatial context, and exposure to stressors contribute meaningfully to variability in clinical presentation and persistence.^5,6^ Despite this, genetic and environmental domains are often modeled separately or combined in simple ways, with environmental variables added alongside genetic predictors rather than incorporated into a unified model.^7^ Such approaches limit our ability to model and understand how different sources of liability compete, complement, or substitute for one another within predictive frameworks.

Efforts to incorporate biological annotation into genetic prediction models have likewise been largely static. Some approaches include entire annotation categories as fixed feature blocks.^8,9^ Other strategies perform pre-model filtering based on external enrichment results or biological priors, selecting annotation subsets before model training begins.^10^ Feature ranking methods such as minimum redundancy–maximum relevance (mRMR)^11^ or LASSO-based selection may iteratively prioritize predefined features, but these approaches generally treat all features separately. In this setting, each SNP or annotation is treated as an independent predictor, without modeling relationships among features during selection. In most cases, decisions like SNP inclusion thresholds, annotation depth, and environmental domains are specified in advance, and model architecture is optimized independently of feature selection. As a result, variant inclusion, biological context, environmental domains, and network structure rarely compete within a single unified objective function.

Like prior work ^12^ we address the problem of predicting clinician diagnoses of ADHD using data collected before or at the same time as the clinician assessment. Although such models are unlikely to replace clinician diagnoses, they could be used to screen patients for ADHD or to help clinicians diagnose complex or uncertain cases. That such models can achieve good levels of accuracy is seen in a report from the Swedish registries showing that clinical, demographic, educational and forensic data predicted ADHD with an area under the curve (AUC) statistic of 0.75. That work used a deep neural network but did not incorporate genetic data.

We hypothesized that a joint optimization framework might overcome these limitations. If GWAS p-value thresholds, annotation depth, environmental block inclusion, and architectural parameters were treated as tunable hyperparameters, the optimization process itself could be informative. Selection patterns across high-performing models could reveal which biological annotations consistently improve prediction, whether deeper annotation representations add meaningful information, and whether different domains contribute shared or distinct predictive signals. In this way, optimization might function not only as a predictive tuning strategy, but also as a data-driven prioritization method for interrogating ADHD risk architecture.

Multi-modal classification models have been shown to be more predictive than unimodal models in a variety of clinical settings.^13^ They are particularly relevant for clinical decision support because clinical judgements, like those made in diagnosing ADHD, are similarly made by integrating heterogenous sources of information. Unlike generative language models, which can produce fluent but unsupported output with little information on confidence,^14^ classification models can provide quantified risk estimates and machine learning explainability^15^ methods help identify the reasoning behind each prediction. Understanding which features lead to model predictions could guide clinicians towards patient characteristics that might otherwise be missed and could help researchers prioritize future research directions.

Here, we develop and evaluate a multi-modal deep learning modeling approach applied to the Oregon ADHD-1000 cohort that integrates SNP-level genotype data with structured biological annotation, polygenic risk and resilience scores, and multiple environmental domains. We treat GWAS p-value thresholding, annotation depth, environmental inclusion, and model architecture as jointly optimized components within a unified hyperparameter search. We compare the performance of single-domain and multi-domain models and examine hyperparameter selection and feature importance patterns to ascertain which genetic annotations and environmental domains most consistently contribute to ADHD prediction. Through this approach, we aim to advance both predictive modeling, clinical utility of prediction models, and empirical prioritization of risk-relevant biological and environmental features in ADHD.

## Methods

### Overview

We built a modular, multi-modal deep learning pipeline to predict ADHD case-control status from diverse input features, including individual genomic features, polygenic scores, demographics, parenting and family conflict, stress and trauma, geocoded measures, cognitive task measures, temperament factor scores, and missingness indicators. We used a single optimization framework to tune model architecture while also allowing the data to determine whether to include the genomic block, which GWAS p-value threshold to use for SNP inclusion, the depth of selected annotation categories, and which additional feature blocks to include.

### Input data and preprocessing

We analyzed participants from the Oregon ADHD-1000^16^ cohort with previously generated genotype data. The participant demographics for Oregon ADHD-1000 are presented in Table 1. We used Year 1 ADHD status as the classification label. ADHD status was established by a child psychiatrist and child psychologist who independently diagnosed participants based on parent and teacher rating scales, semi-structured clinical interviews, a short form of the WISC-IV to estimate IQ, achievement testing screeners, the Stroop and Trailmaking cognitive tests, and clinician and psychometrician written observations. We split participants randomly into training (70%), validation (15%), and testing (15%) subsets. We calculated associations and trained models using the training subset (n = 537; 343 cases and 194 controls), optimized hyperparameters using the validation subset (n = 114; 70 cases and 44 controls), and tested performance using the test subset (n = 114; 64 cases and 50 controls), We organized baseline measures into feature blocks to support block-wise inclusion during optimization, including 35 demographic features, 25 parenting and family conflict features, 64 stress and trauma features, 7 geocoded measures, 17 cognitive task measures, 12 temperament factor scores, and 2 polygenic score features. The full list of features can be found in Supplementary Table 1. We imputed missing continuous variables with the median and missing categorical variables with the mode, then one-hot encoded categorical variables and z-score scaled continuous variables. To avoid issues of data leakage, the scaling of continuous variables in validation and test sets was done using the means and standard deviations calculated in the training subset. We also created missingness indicators for each feature and treated them as a dedicated feature block.

**Table 1.** Demographic information for the Oregon ADHD-1000 data set.

|  |  |  |
| --- | --- | --- |
| % Female | 39% |  |
| Mean Age<sup>a</sup> (SD) | 9.4 years (1.6) |  |
| Mean Family Income<sup>b</sup> (SD) | 4.5 (2.0) |  |
| Race / Ethnicity | Not Hispanic /<br>Latino | Hispanic /<br>Latino |
| American Indian / Alaska Native | 3 (0.2%) | 2 (0.1%) |
| Asian | 30 (2.1%) | 1 (0.07%) |
| Native Hawaiian / Pacific Islander | 5 (0.3%) | 2 (0.1%) |
| Black / African American | 38 (2.6%) | 3 (0.2%) |
| White | 1,148 (79.2%) | 54 (3.7%) |
| More than One Race | 123 (8.5%) | 34 (2.3%) |
| Unknown / Not Reported | 5 (0.3%) | 2 (0.1%) |

<sup>a</sup> Age at baseline
<sup>b</sup> Family income measures are based on the following scale: 1 = less than \$25,000, 2 = \$25,000 - \$35,000, 3 = \$35,000 - \$50,000, 4 = \$50,000 - \$75,000, 6 = \$100,000 - \$130,000, 7 = \$130,000 - \$150,000, 8 = more than \$150,000

We paired SNPs with a structured annotation matrix to provide biological context for convolutional neural network (CNN) modeling. We created an annotation matrix with three components: 1) the functional genomic annotations used in SBayesRC;^9^ 2) an annotation block with information on whether each SNP was present in the 100 gene ontology gene sets (collection c5.all.v7.1)^17^ that were most associated with ADHD in the training subset based on MAGMA^18^ analysis; and 3) an annotation block containing risk estimates from GWASs of disorders genetically correlated with ADHD based on the GWAS Atlas. We have previously found that inclusion of both gene set and correlated disorder information improves prediction of ADHD.^19^

We generated ancestry principal components (PCs) from a separate SNP set using PLINK^20^. We created an ancestry-focused SNP set, applied linkage disequilibrium (LD) pruning, computed and standardized the top 10 PCs, and merged them into the train, validation, and test input files.

We used these 10 PCs as a secondary prediction target in the adversarial component of the model. We have previously shown that this effectively removes the models ability to predict ancestry,^21^ and because the adversarial task and main task share all layers, ancestry information cannot be used for the main task. This helps mitigate the risks of population stratification and prevents conflating genetics associated with ancestry and genetics associated with disease.

### Model architecture, optimization, and evaluation

Figure 1 presents an overview of our model architecture. If the genomic data block was selected for model inclusion, we used the SNP input alongside the annotation matrix into 1 – 3 genomic convolutional blocks, which we have described previously.^21^ We flattened the convolutional output and concatenated it with any included environmental blocks. We then passed the combined features through 1 - 3 fully connected layers followed by an output layer for ADHD prediction.

**Figure 1:**
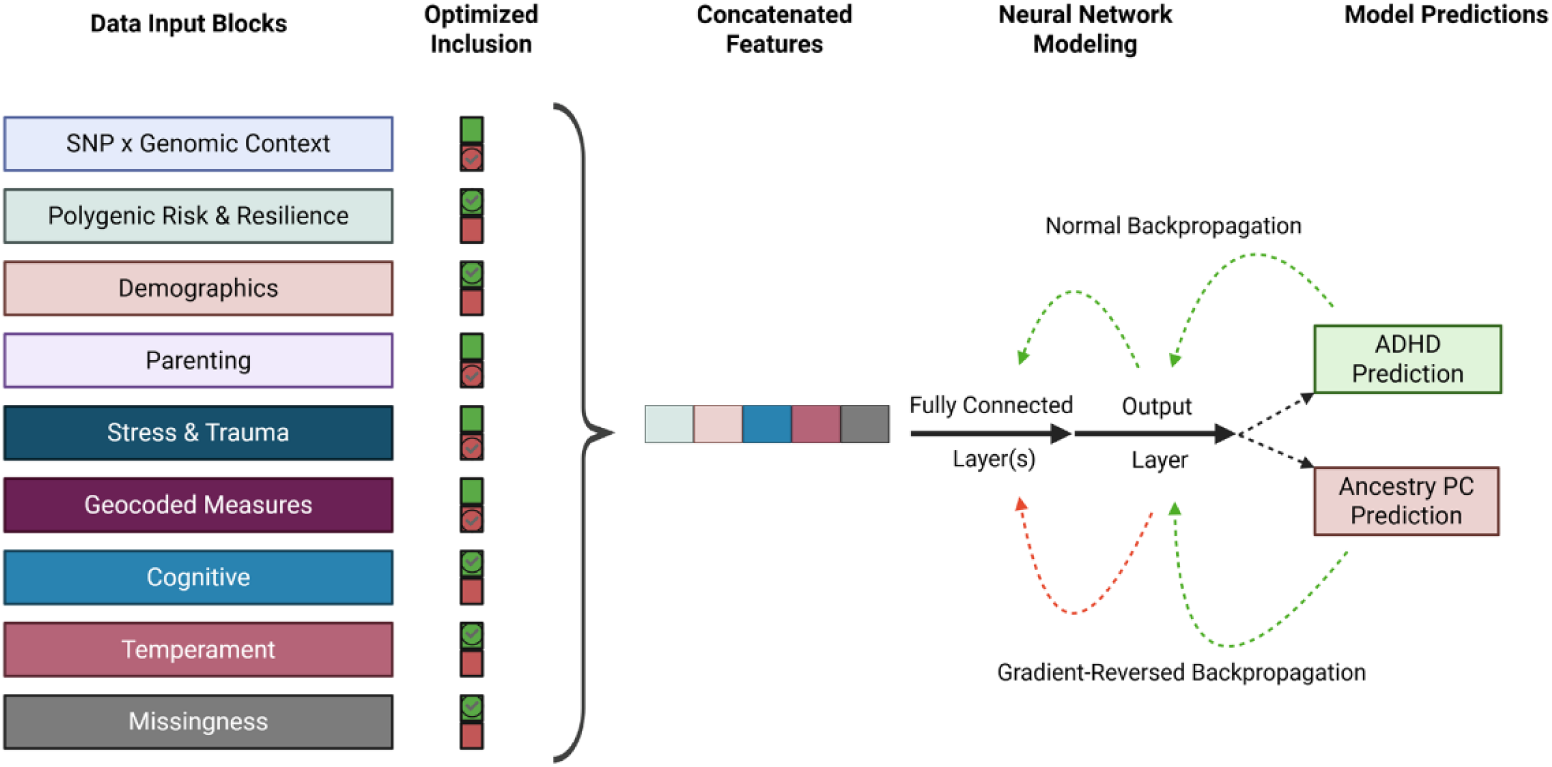
Model Architecture Diagram. Overview of the multi-modal deep learning framework used to predict ADHD case-control status. Hyperparameter optimization determined inclusion of several multi-modal feature blocks including output from a genomic convolutional neural network model, polygenic scores, demographics, parenting and family conflict, stress and trauma, geocoded measures, cognitive task measures, temperament factor scores, and missingness indicators. The selected feature blocks were combined and used as input into a fully connected neural network with a primary ADHD prediction task and a secondary adversarial task predicting ancestry principal components through a gradient reversal layer. Green arrows indicate normal backpropagation and red arrows indicate gradient-reversed backpropagation.

We added an additional output layer for a secondary task to predict the 10 ancestry PCs and placed a gradient reversal layer before the output layer of this task. This encouraged the network to learn features that supported ADHD prediction while "unlearning" information predictive of ancestry structure. We treated the gradient reversal weight as a tunable hyperparameter.

We optimized hyperparameters with Optuna^22^ using validation AUC as the objective. The search space included architectural parameters (number of convolutional blocks, filters, kernel widths, pooling widths, number of dense layers, dense units, dropout rates, learning rate, epochs), genomic selection parameters (whether to include the genomic block, GWAS p-value threshold for SNP inclusion, and the number of gene set and correlated-GWAS annotation rows), the gradient reversal weight for PC unlearning, and Boolean inclusion flags for the additional feature blocks. Hyperparameter ranges are shown in Supplementary Table 2.

We trained models with Adam optimization using a joint loss that summed binary cross-entropy for ADHD prediction and mean squared error for PC prediction. For each Optuna trial, we evaluated performance on the validation set using AUC. To reduce sensitivity to random initialization and optimization instability, we repeated training within each trial across ten random seeds and used mean validation AUC as our optimization target. After selecting the best hyperparameter configuration, we evaluated the resulting model on a held-out test set. We estimated 95% confidence intervals using bootstrap resampling.

We evaluated the performance of each multi-modal model using several additional approaches, including precision-recall curves, model calibration analyses, and decision curve analysis (DCA). Precision-recall curves were created to illustrate the tradeoff between positive predictive value (precision) and sensitivity (recall) when using different case/control thresholds. Model calibration analyses tested whether each model’s predicted probabilities matched observed outcomes. We calculated Brier scores, which measure the average squared difference between predicted probability and true outcome, and generated calibration plots for each model. In a related analysis, we tested whether our models were less confident about predictions that were incorrect. For this analysis, we stratified prediction certainty, defined as the distance of a prediction from 0.5, based on prediction correctness and performed Welch’s t-test to compare the mean prediction certainty in correct and incorrect predictions. We used DCA to test whether each model would be useful for clinical decision making. DCA estimates net benefit of using a model to take some clinical action across different threshold probabilities compared to action for everyone and action for no one. Threshold probabilities represent the risk level threshold (i.e., the likelihood that the patient has the disorder) at which the clinician and patient would choose to take some action. We estimated confidence intervals for the precision-recall curves, DCA, and calibration curves using 1,000 bootstrap replicates.

### Feature importance analysis

After model optimization, we estimated feature importance using integrated gradients, which quantified the contribution of each input feature to the model prediction relative to a reference baseline defined by the mean feature values in the validation subset. We summarized these attributions at both the individual feature level and the block level, and we also generated person-level feature importance outputs to support individualized explainability and interpretation of model predictions. We applied these analyses to grouped models fit with and without temperament features. This approach allowed us to evaluate both the relative contribution of broad feature domains and the specific variables driving model performance under alternative feature-set definitions. We used 1,000 bootstrap samples to estimate the confidence intervals of each feature importance score.

To evaluate the empirical significance of these feature importance results, we generated a null distribution by permuting the training labels, retraining the model with the same optimized hyperparameters, and recomputing integrated gradients on the same test inputs. We repeated this procedure 10,000 times and calculated empirical p-values for each block and feature as the proportion of null replicates with absolute attribution values greater than or equal to the observed value. For individual features, we also applied false discovery rate (FDR) correction to the resulting empirical p-values.

### Alternative classification labeling analysis

While the diagnostic team that produced the ADHD diagnostic labels was independent from the data collection teams, there was overlap in the information used by the diagnostic team and our models. These overlaps included the Stroop and Trailmaking cognitive tests as well as measures of attention and impulsivity. While the TMCQ data used in the models were not used in the diagnostic decision-making, the content of some TMCQ questions is similar to the parent-reported symptom rating scales used by the diagnostic team. This introduced a potential source of circularity, in that the model was asked to predict a label derived from information similar to its own inputs. To assess the magnitude of this potential problem, we used KSADS-based ADHD diagnostic labels as well. These models contained an additional output task that aimed to predict the diagnostic team-based ADHD label, but that task was disconnected from the rest of the model and therefore had no effect on model training beyond the task-specific output layer. This additional task was used to compare classification performance across ADHD labeling approaches. The methods used to build these models were otherwise identical to the models built to predict diagnostic team-derived ADHD labels.

## Results

### Performance in classifying diagnostic team-based ADHD diagnosis

We first evaluated the more inclusive models that empirically selected which feature blocks to include. The best performing model in the analysis that included temperament features achieved an AUC of 0.97 (95% CI: 0.93 - 0.99), an accuracy of 0.91 (95%CI: 0.76 - 0.97), a sensitivity of 0.94 (95% CI: 0.88 - 0.99), and a specificity of 0.88 (95% CI: 0.79 - 0.97) in the test subset. This high level of accuracy is not surprising because two of the temperament factors assess inattention and impulsivity.

Hyperparameter optimization selected the temperament, demographics, cognitive, stress and trauma, and geocoded blocks for final model inclusion. The best performing model in the analysis that excluded temperament features achieved an AUC of 0.75 (95% CI: 0.65 - 0.83), an accuracy of 0.66 (95% CI: 0.57 - 0.75), a sensitivity of 0.78 (95% CI: 0.67 - 0.87), and a specificity of 0.52 (95% CI: 0.38 - 0.66) in the test subset. Hyperparameter optimization selected the demographics, cognitive, stress and trauma, geocoded, and polygenic score blocks for model inclusion. The hyperparameters selected for each model are in Supplementary Table 3.

Precision-recall curves for both multi-modal models are shown in Supplementary Figure 1. The model with temperament had an average precision (AP) of 0.97 (95% CI: 0.94 – 1.00) while the model without temperament had an AP of 0.82 (95% CI: 0.73 – 0.89). The calibration plots for both models (Supplementary Figure 2) did not show evidence of miscalibration, as the confidence intervals around the observed event rates overlapped the ideal calibration line across calibration bins. The model with temperament had a Brier score of 0.06 (95% CI: 0.03 – 0.09) and Brier skill score of 0.70 while the model without temperament had a Brier score of 0.22 (95% CI: 0.18 - 0.27) and Brier skill score of 0.10. In our prediction certainty analysis, correct predictions were significantly more certain compared to incorrect predictions (Welch’s t-test p: 0.01) with a mean prediction distance from null of 0.43 in correct predictions compared to 0.24 in incorrect predictions. DCA indicated that the model that included temperament had a positive net benefit compared to clinical action for all or clinical action for none at probability thresholds from 0.22 to 0.89. The model that did not include temperament was no better than action for all or action for none at any probability threshold. Supplementary Figure 3 shows DCA plots with confidence intervals.

### Feature importance in multi-modal models

Figure 2 shows the results for the model fit with temperament features, and Supplementary Figure 4 shows the results for the model fit without temperament features. In each figure, panel A summarizes block-level feature importance, and panel B shows the distribution of block-level feature importance across people.

**Figure 2.**
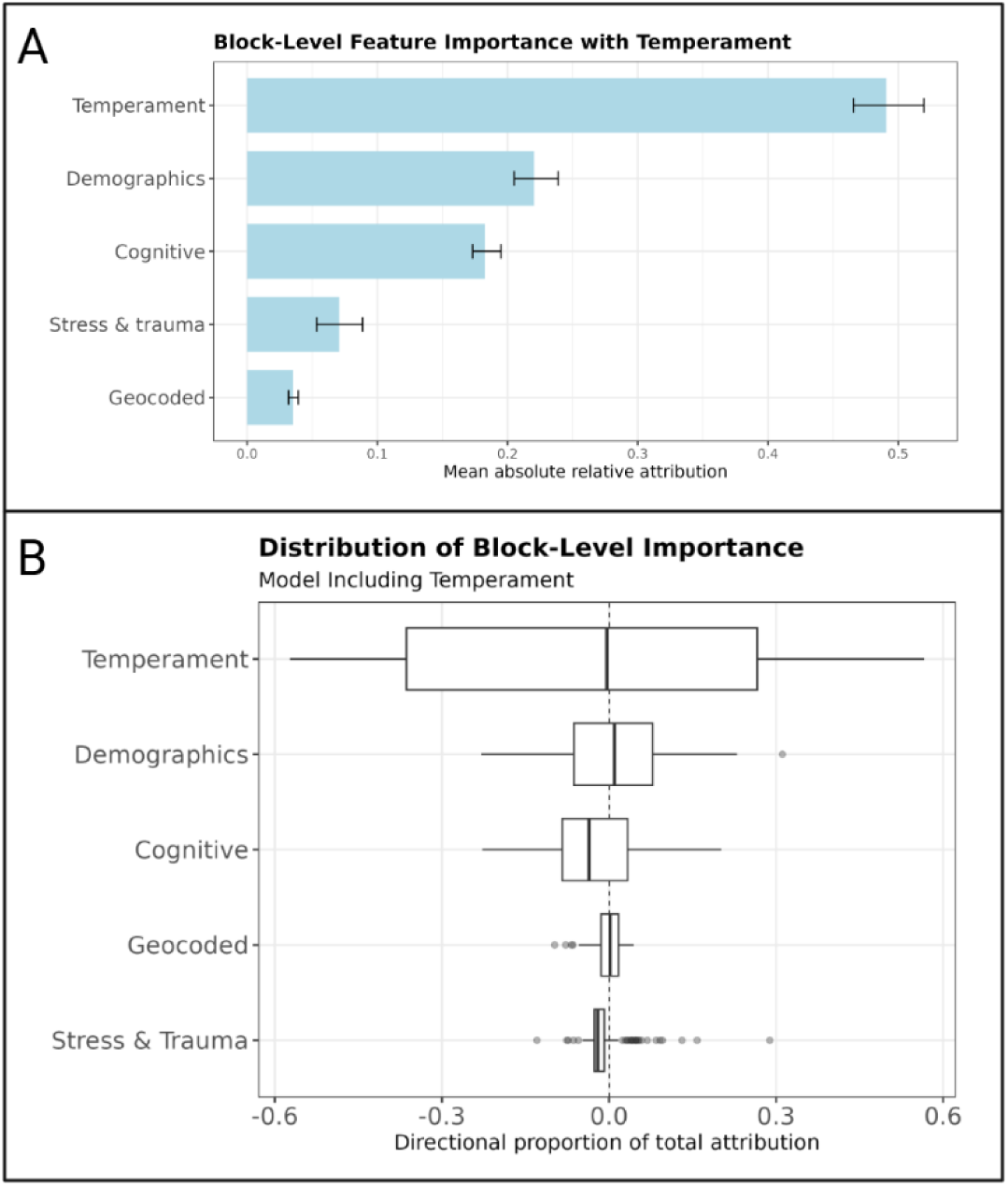
Feature Importance Results: Including Temperament. Feature importance results for the best-performing jointly optimized model that allowed inclusion of temperament factor scores with the task of classifying diagnostic team ADHD diagnosis. Panel A shows block-level integrated gradient attribution results, summarizing the overall contribution of each included feature domain to ADHD prediction. Panel B shows a boxplot of the distribution of block-level feature importance results for the most influential variables contributing to ADHD prediction across participants in the test subset. Feature importance values were calculated relative to a reference baseline defined by the mean feature values in the validation subset, and 95% confidence intervals were estimated using 1,000 bootstrap samples.

In the temperament-inclusive model, the block-level results highlighted temperament, demographics, and cognitive feature blocks. At the individual-feature level, the most influential features were attention and impulsivity temperament factor scores, followed by age of child at screening, activity temperament factor score, the color naming condition of the Stroop task, and sex. In total, the feature importance scores of 66 features had an FDR corrected empirical p-value < 0.05. Individual variable definitions are in Supplementary Table 4. Complete block-level and feature-level feature importance results are in Supplementary Tables 5 and 6.

In the model without temperament features, the block-level results highlighted demographic and cognitive feature blocks. At the individual-feature level, the most influential feature was age of child at screening, followed by discrimination results in a continuous performance test, sex, average stop-signal reaction time results, a parenting stress index indicating whether the child had trouble with teachers at school, and living arrangement, among others. The feature importance scores of 14 features had an FDR corrected empirical p-value < 0.05. This included 7 demographic features, 4 stress and trauma features, 1 cognitive feature, and polygenic risk and resilience scores. Feature importance results for this model are available in Supplementary Tables 7 and 8.

Figures 3 and 4 present four random examples from the test subset of individualized feature importance results measured by the proportion of each individual’s total attribution explained by a feature. Figure 3 shows output examples from block-level individualized feature importance analyses and Figure 4 shows output examples from feature-level individualized feature importance analyses. Supplementary Figure 5 shows a scatter plot of the block-level feature attribution by prediction confidence percentile of all individuals in the test subset.

**Figure 3.**
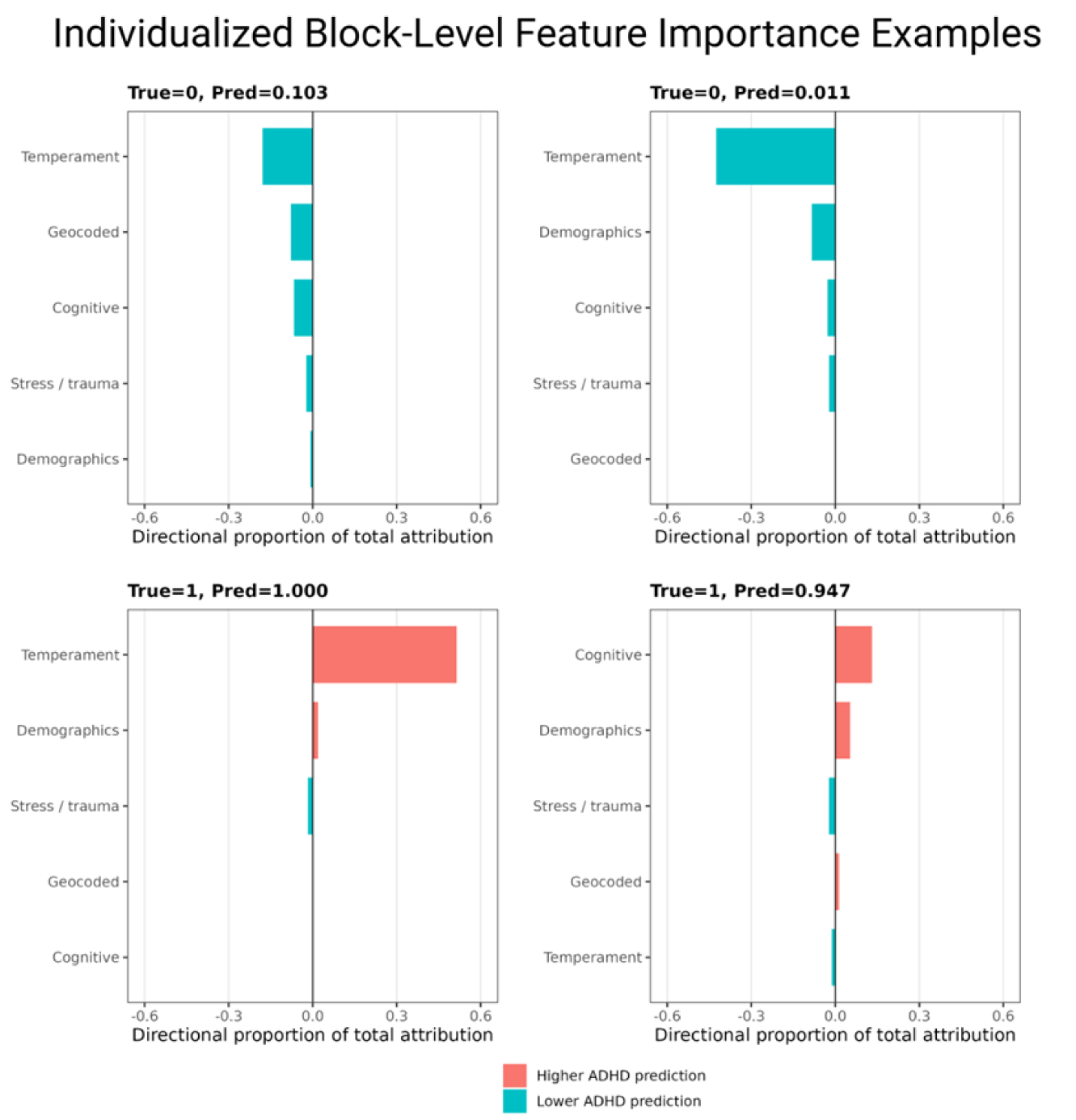
Individualized Block-Level Feature Importance Examples. Random examples of person-level block-wise feature importance outputs from the optimized models. Each panel shows the contribution of broad feature domains to the predicted ADHD risk for one individual participant, illustrating how the relative influence of domains can vary across people. Positive and negative attributions indicate whether a feature block increased or decreased the model’s predicted ADHD risk for that individual. True represents the participants diagnosis label (0 = control, 1 = case) and Pred represents the model’s label prediction.

**Figure 4.**
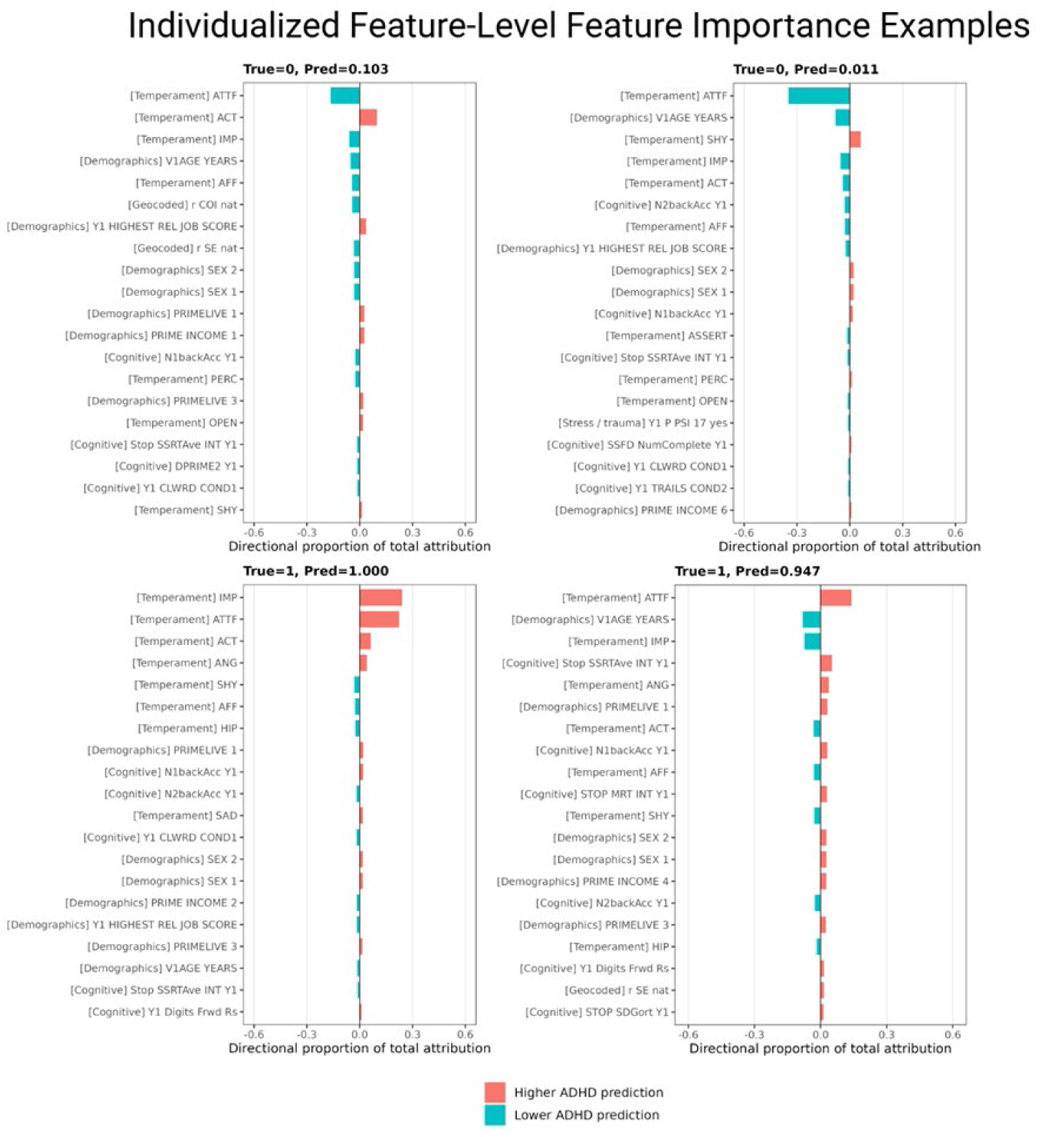
Individualized Feature-Level Feature Importance Examples. Examples of person-level feature-specific importance outputs with the same individuals as Figure 3. Each panel shows the contribution of individual variables to the predicted ADHD risk for one participant, illustrating the heterogeneity of model explanation profiles across individuals. Positive and negative attributions indicate whether a given feature increased or decreased the model’s predicted ADHD risk for that person. True represents the participants diagnosis label (0 = control, 1 = case) and Pred represents the model’s label prediction.

Supplementary Figure 6 presents a scatterplot of the within-block feature attribution discordance, defined as the range between the most negative and most positive feature attribution within each block, by prediction confidence percentile of all individuals in the test subset. Figure 5 illustrates the potential for conflicting or contradictory information between attention and impulsivity features in two ways: Panel A plots the directional proportional attention attribution against the impulsivity attribution for each person in the test subset. Panel B is a scatter plot of the discordance between attention and impulsivity attributions by prediction confidence of all individuals in the test subset. Among cases, attention/impulsivity discordance decreased significantly as prediction confidence increased (r = -0.53, p = 7.2 x 10^-6^), suggesting that lower-confidence predictions had more conflicting evidence. Among controls, there was no significant correlation between attention/impulsivity discordance and prediction confidence (r = -0.13, p = 0.34).

**Figure 5.**
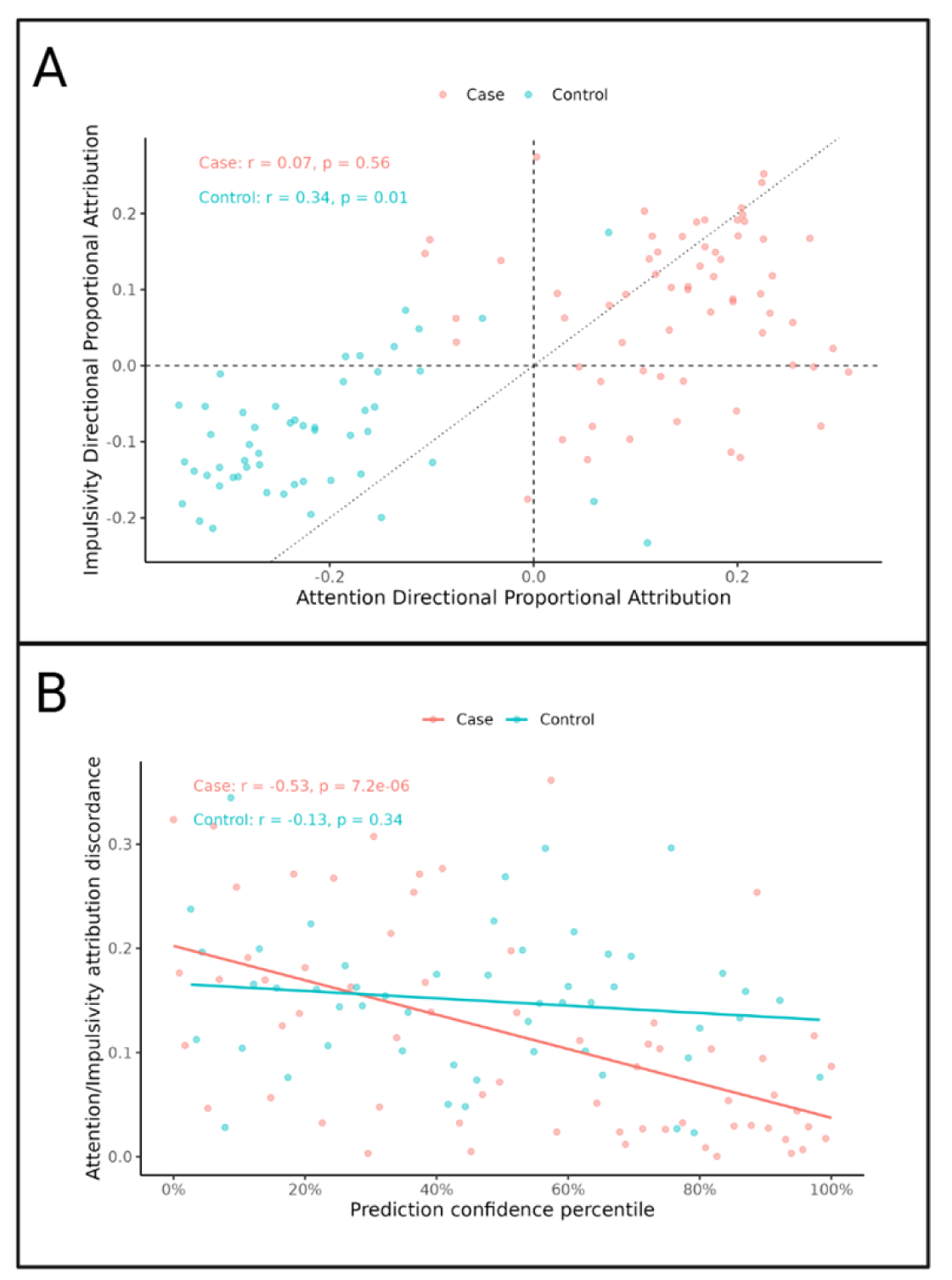
Attention/Impulsivity Attribution Discordance. Panel A presents a scatter plot comparing the directional proportional attributions for the attention temperament feature (ATTF) and impulsivity temperament feature (IMP). Positive values indicate evidence pushing the model towards ADHD case status and negative values indicate evidence pushing the model towards control status. The upper-left and lower-right quadrants represent contradictory temperament evidence, where ATTF and IMP push the prediction in opposite directions. Panel B presents a scatter plot showing the relationship between prediction confidence percentile and attribution discordance ATTF and IMP. Attribution discordance was defined as the absolute difference between the directional proportional attributions for ATTF and IMP, with larger values indicating greater disagreement between attention- and impulsivity-related evidence. Among ADHD cases, discordance significantly decreased as prediction confidence increased (r = -0.53, p = 7.2 x 10^-6^), suggesting that lower-confidence case predictions had more conflicting attention/impulsivity evidence. This pattern was not significant among controls (r = 0.13, p = 0.34).

### Performance of block-specific models

To further evaluate the independent predictive value of each feature domain, we trained separate models for each block. Performance metrics for these block-specific models are summarized in Supplementary Table 9. Across individual blocks, AUCs ranged from 0.54 (geocoded measures) to 0.99 (temperament), sensitivities ranged from 0.30 (stress and trauma) to 0.98 (geocoded measures), and specificities ranged from 0.06 (geocoded measures) to 0.87 (temperament).

### Performance in classifying K-SADS-based ADHD diagnosis

In the multi-modal models trained to classify based on K-SADS diagnoses of ADHD, the best model included temperament, polygenic risk, and geocoded measures. That model had an AUC of 0.95 (95% CI: 0.92 – 0.98) in the task of predicting K-SADS ADHD diagnoses and an AUC of 0.99 (95% CI: 0.97 – 1.00) in the task of predicting diagnostic team-based ADHD diagnoses. The best model that excluded temperament used the polygenic risk, demographics, parenting, and cognitive blocks and had an AUC of 0.70 (95% CI: 0.60 – 0.78) in the K-SADS-based ADHD task and an AUC of 0.68 (95% CI: 0.68 – 0.77) in the diagnostic team-based ADHD task.

## Discussion

Our results suggest that, in the absence of highly predictive features, models that use multiple data types may improve both predictive performance and the combination of multi-modal modeling and explainability methods can aid model interpretation even in the presence of highly predictive features. The high performance of the multi-modal and individual-block models that included temperament features was expected and fits with the nature of the available data. Two of the temperament factor scores are closely related to the behavioral dimensions used to identify ADHD, so strong predictive performance is not surprising, even though the temperament scores and diagnostic labels were derived from different assessment teams.^23,24^ Prior work has shown that, while not as consistent as statistical prediction, clinical decisions are generally consistent when based on the same core behavioral information.^25^ Our results are in line with that expectation. For this reason, we also trained a model without temperament features to give other data types an opportunity to contribute to prediction rather than being overshadowed by temperament scores. Although performance was lower in the model without temperament, it still reached a level that suggests meaningful predictive value (AUC of 0.75).

Regarding the temperament scores, the inattention and impulsivity factors were the most important predictors in our models. These features overlap closely with the information clinicians use when making ADHD diagnoses, including parent and teacher reports and semi-structured interview data focused on attention difficulties and impulsive behavior. In that sense, high classification accuracy in models that include temperament is expected and demonstrate the diagnostic consistency between clinicians and those models. While our temperament only model was not significantly different in performance compared to our multi-modal model, in a clinical setting, where clinicians are already accustomed to focusing on temperament-related information,^26^ a model built from non-temperament variables could serve as a useful helper by providing an estimate of ADHD risk from sources of information that may otherwise receive less emphasis.

The rise in popularity of large language models (LLMs) has spurred research into using artificial intelligence as clinical decision support systems, which has renewed concerns regarding model hallucinations, false-confidence, and lack of interpretability.^14^ Our modeling and analysis approach, though not an LLM, aims to address those issues through model performance metrics and explainability methods. In our temperament inclusive model, the Brier score (0.06), Brier skill score (0.70), and significant confidence difference between correct and incorrect predictions suggest that the model is well-calibrated, meaning that the confidence in which the model makes predictions is in line with the accuracy of those predictions. Our feature importance analyses provide insights into how our model calculated each decision.

Our findings highlight the value of feature importance analyses for both research and clinical contexts. From a research perspective, feature importance identifies which domains and variables warrant greater attention in future studies and which types of data may be most useful to collect. Clinically, knowing which model inputs contributed to a prediction could help direct attention toward factors outside the usual focus of an ADHD evaluation and could support decisions about diagnosis, triage, or follow-up assessment. With sufficiently high diagnostic consistency between clinicians and a clinical decision support model, discordance between clinician and model diagnosis could promote the clinician to consider the features the model found most important.

For example, in the bottom right patient example shown in Figures 4 and 5, we show a patient who was diagnosed with ADHD by the ADHD-1000 diagnostic team and predicted to have ADHD by our model. According to the model’s block-level feature importance, the decision is based largely on cognitive features, whereas typically temperament features were most important. In a clinical setting, a clinician might lean towards not diagnosing this patient with ADHD, citing their lack of impulsivity features. However, upon considering that 1) the model was relatively confident in its ADHD prediction, 2) the block-level results that cognitive features were more predictive in this patient, and 3) the feature-level results considered contradictory temperament features—low impulsivity, which is predictive of lower ADHD risk, and low attention, which is predictive of higher ADHD risk—but predicted the patient to have ADHD using the supporting evidence of several cognitive tests, the clinician might take a closer look at the cognitive test results. The attention/impulsivity attribution discordance results presented in Figure 5 show that attention and impulsivity features often offer differing and potentially contradictory evidence for ADHD prediction. This suggests that scenarios like the one described here, where additional supporting evidence would be useful in overcoming an unclear or contradictory clinical picture, are relatively common. Ultimately, this process could lead to more informed, but still clinician-led, clinical decisions, especially for patients who are difficult to diagnose based on the clinical interview.

Our results also illustrate the trade-off between the number of features and the number of hyperparameters with a finite sample size. In these analyses, hyperparameter optimization did not select the genomic CNN portion of the model, even though we have previously seen value from this component in other work.^21^ The present dataset is substantially smaller, however, and it also includes additional genetic and non-genetic features that may overlap with part of the predictive information carried by SNP-level data. Under those conditions, the model appears to have favored a simpler architecture that did not require learning the large number of parameters associated with the genomic CNN. The same logic likely applies to the exclusion of other feature blocks. Their omission should not be interpreted as evidence that they contain no useful or predictive information, but rather as evidence that, in this dataset, the gain in information did not justify the added model complexity. Larger datasets may shift these trade-offs and support inclusion of blocks that were not selected here.

Several limitations should be considered when interpreting these findings. First, the sample size was modest for a machine learning study that aimed to compare multiple feature domains and allow flexible block inclusion. Our limited sample size limited our ability to statistically compare models, given the large confidence intervals our small test subset produced. However, the performance of our models, including those without temperament factor scores, is notable given the limited training size. Second, the high performance of the temperament-inclusive model should be interpreted in light of the rater and content overlap between the temperament measures and the behavioral information clinicians used to formulate the diagnoses of ADHD. More broadly, both diagnosis and several of the strongest predictors relied on parent- and teacher-reported information, which would increase circularity. Our alternate models that classified K-SADS ADHD diagnosis had similar performance and were less circular since the exact features used in K-SADS diagnosis and those models were entirely separate. However, it could be argued that some degree of circularity is present in any model that uses clinical features to predict a clinical diagnosis due to the inevitable overlap in clinical information. Third, all models were developed and tested within a single cohort, and the selected architecture and feature importance patterns may therefore reflect characteristics of this particular sample. The depth of the information available and multi-modal nature of the data we used allowed us to test our multi-modal, optimizable approach, but complicates harmonization with external data. Finally, feature importance results indicate which variables contributed to model predictions in this dataset, but they should not be interpreted as causal effects or as proof that these variables would generalize in the same way across other populations.

In summary, these results suggest that even though including data beyond temperament did not improve ADHD classification performance, there is value in multi-modal modeling. We show that explainable machine learning approaches can provide information beyond overall model performance alone. The contrast between the temperament-inclusive and temperament-excluded models highlights both the strength of clinically proximal measures and the potential value of broader supporting features when those measures are not available. The feature importance analyses further show how machine learning models can identify both broad domains and individual variables that contribute to prediction, including at the level of individual patients.

Together, these findings suggest that multi-modal, explainable risk models may be useful for both future research on ADHD liability and the development of clinical tools that support diagnostic decision making and patient triage.

## Scientific Ethical Approval

The SUNY Upstate Medical University IRB determined that this project did not meet the criteria for human subject research.

## Data Availability

Summary-level results generated from the analyses reported in this manuscript are available from the corresponding author upon reasonable request.

## Acknowledgements

The authors thanks and acknowledge Dr. Joel Nigg and his team at the Steven J. Sharp Center for Mental Health Innovation at Oregon Health & Science University for the development of the Oregon ADHD-1000 cohort and samples.

Stephen V. Faraone, PhD, served as the statistical expert for this research.

This research was supported by NIH/NIMH grant U01MH135970. Dr. Barnett’s research over the past two years has been supported by European Union’s Horizon 2020 research and innovation programme under grant agreement 965381 and NIMH grant R01MH131685. Dr. Mooney’s work was supported by NIMH grant R01MH131685. Dr. Ryabinin’s work was supported by NIMH grant R01MH131685. Dr. Zhang-James receives research support from European Union’s Horizon 2020 research and innovation programme under grant agreement No 965381. She reports grants from Tris Pharma, Inc outside the submitted work. Dr. Faraone’s research is supported by the European Union’s Horizon 2020 research and innovation programme under grant agreement 965381; Patient-Centered Outcomes Research Institute (PCORI); NIH/NIMH grants R01MH131685, 1R01MH130899, U01MH135970, Tris Pharmaceuticals, Supernus Pharmaceuticals and Otsuka Pharmaceuticals OAK-ISS-2023-001796. His continuing medical education programs are supported by The Upstate Foundation, Corium Pharmaceuticals, Collegium Pharmaceuticals and Noven Pharmaceuticals.

## Author Contributions

EJB, MAM, and SVF contributed to study conceptualization. EJB, MAM, YZJ, and PR contributed to analytic design and data preparation. EJB conducted the analyses. EJB, MAM, YZJ, and SVF interpreted the results. EJB drafted the manuscript. MAM, YZJ, PR, and SVF critically reviewed and edited the manuscript. All authors reviewed and approved the final manuscript.

---

**Supplementary Figure 1:**
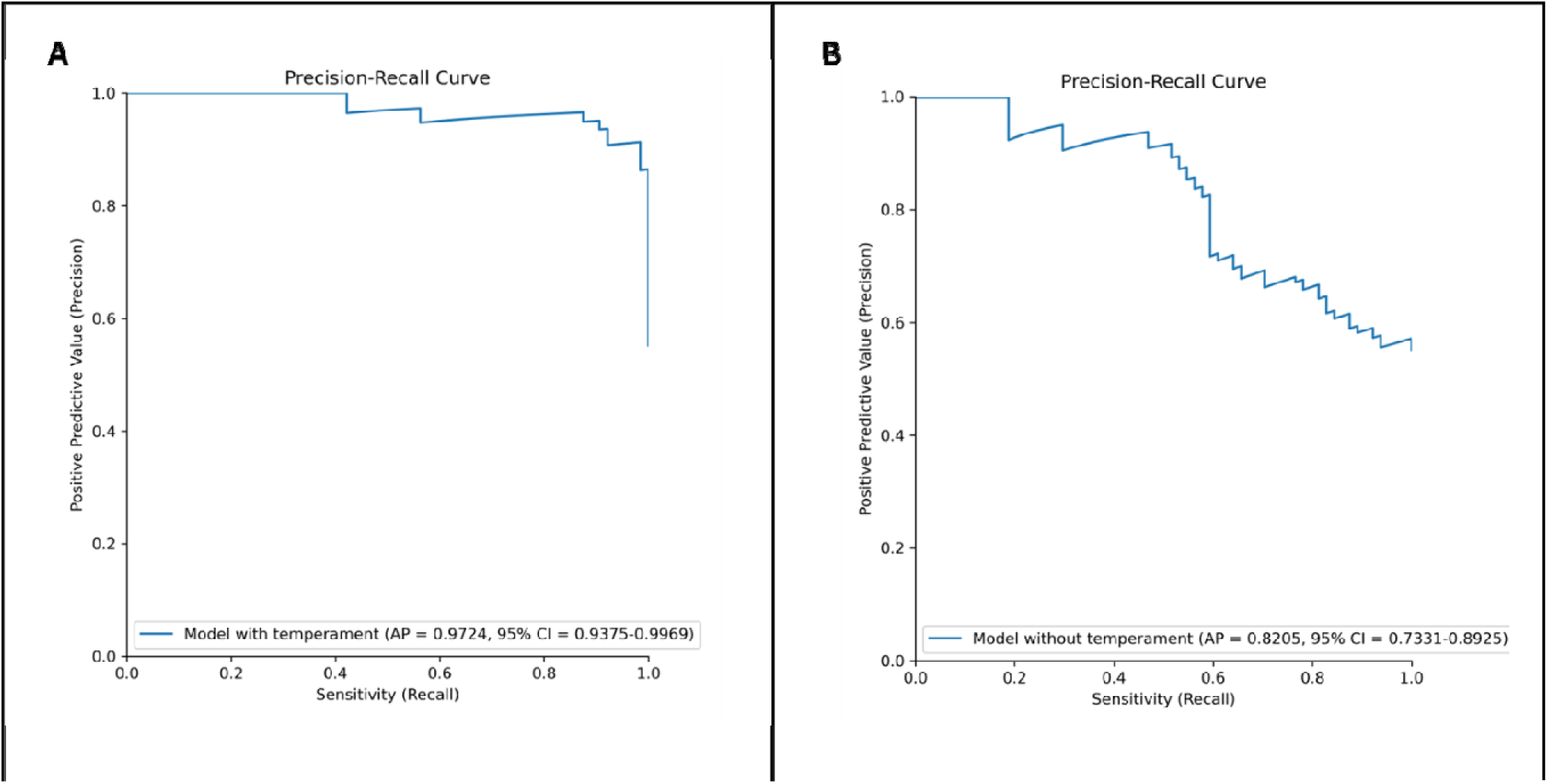
Precision Recall Curves. The precision recall curves for ADHD classification models with and without temperament. Precision-recall curves are shown for the optimized multimodal model including temperament features (left) and the model excluding temperament features (right). The model including temperament had an average precision (AP) of 0.97 while the model without temperament had an AP of 0.82.

**Supplementary Figure 2:**
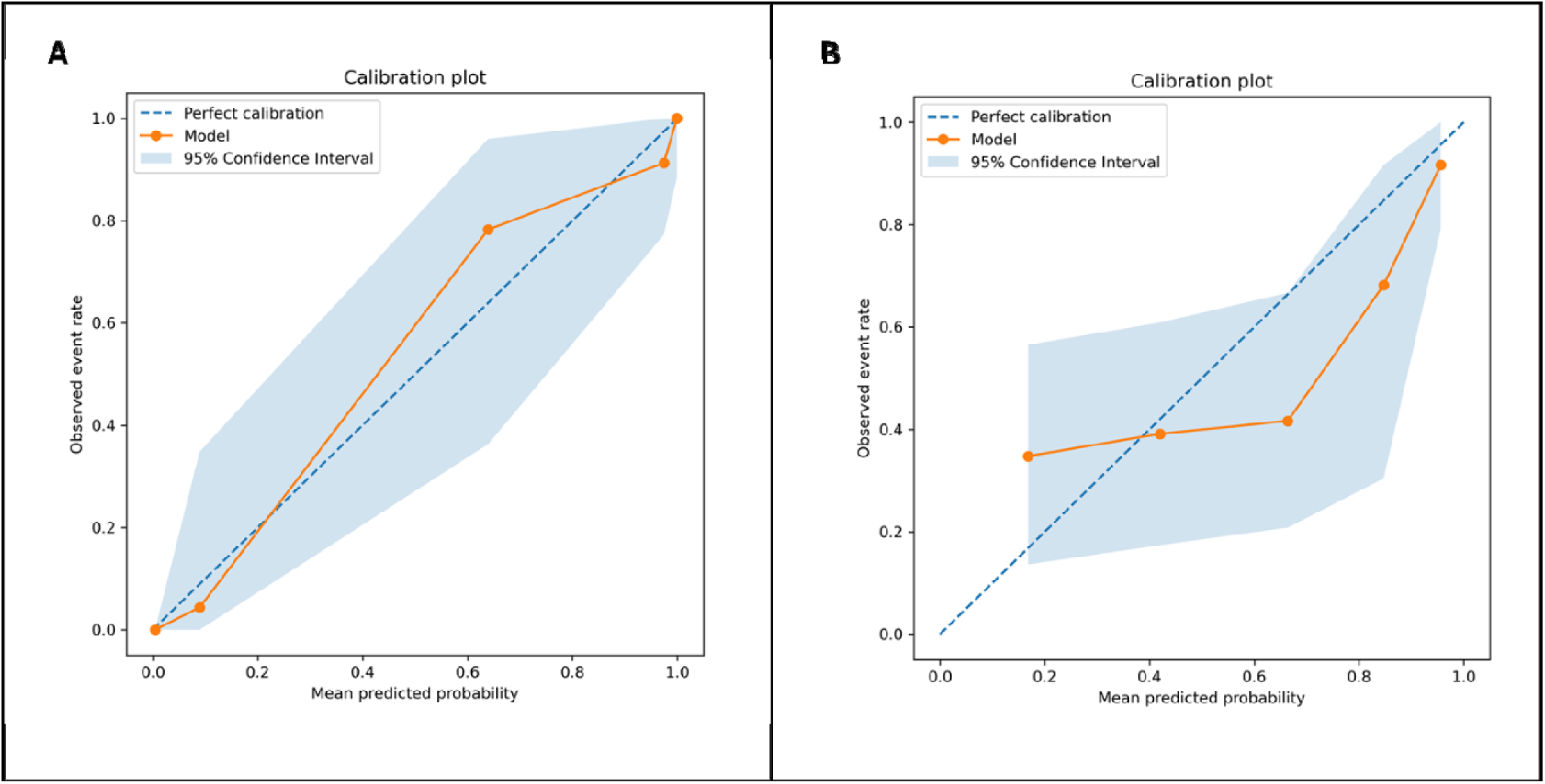
Calibration Plots for ADHD Classification Models. Calibration plots are shown for the optimized multimodal model including temperament features (left) and the model excluding temperament features (right). The dashed line represents perfect calibration, where predicted probabilities match observed event rates. The solid orange line shows the model’s observed calibration across probability bins, and the shaded region indicates the 95% confidence interval. Both models generally tracked the ideal calibration line within the confidence intervals, although the model without temperament showed greater uncertainty and more deviation from perfect calibration across intermediate predicted probabilities.

**Supplementary Figure 3:**
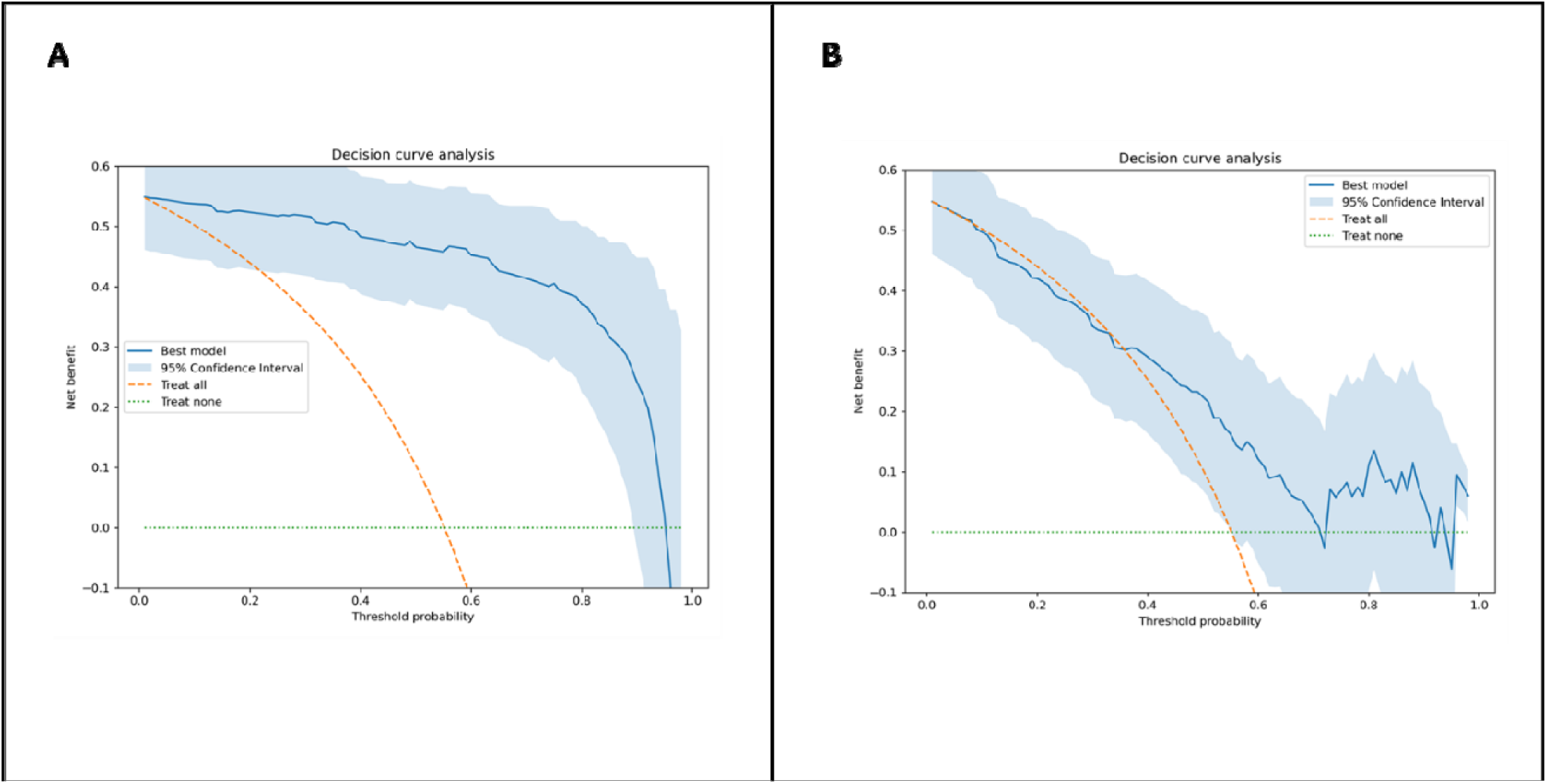
Decision Curve Analyses. Decision curve analyses are shown for the optimized multimodal model including temperament features (left) and the model excluding temperament features (right). Net benefit is plotted across probability thresholds, with the treat-all and treat-none strategies shown as reference lines. The shaded region indicates the 95% confidence interval for model net benefit. The model including temperament showed consistently greater net benefit than the treat-all and treat-none strategies across a broad range of probability thresholds. The model without temperament showed no net benefit after taking confidence intervals into account and showed less stable net benefit at higher threshold probabilities.

**Supplementary Figure 4.**
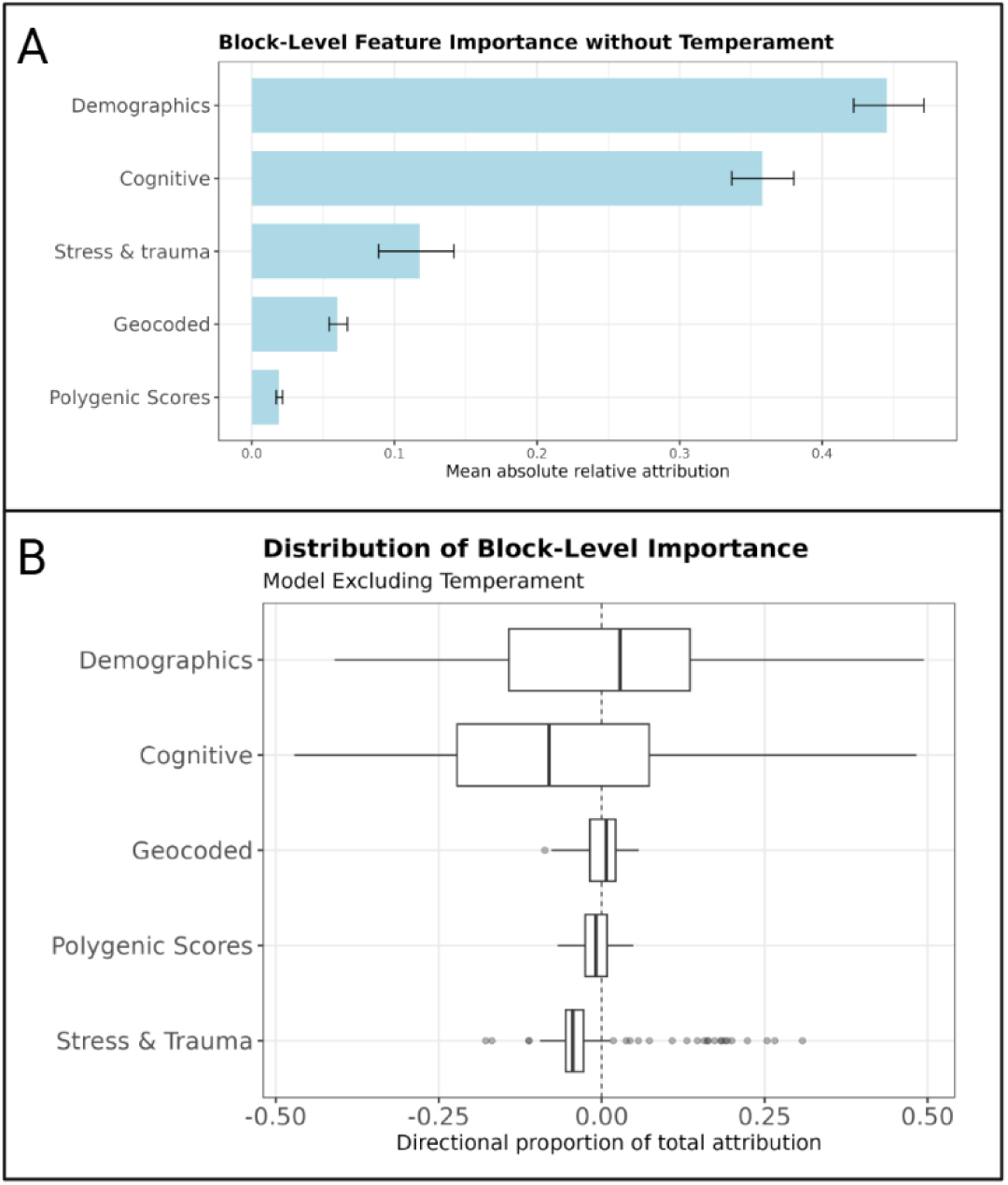
Feature Importance Results: Excluding Temperament. Feature importance results for the best-performing jointly optimized model fit after excluding temperament factor scores with the task of classifying diagnostic team ADHD diagnosis. Panel A shows block-level integrated gradient attribution results for the included feature domains. Panel B shows a boxplot of the distribution of block-level feature importance results for the most influential variables contributing to ADHD prediction across participants in the test subset. Feature importance values were calculated relative to a reference baseline defined by the mean feature values in the validation subset, and 95% confidence intervals were estimated using 1,000 bootstrap samples.

**Supplementary Table 1:**
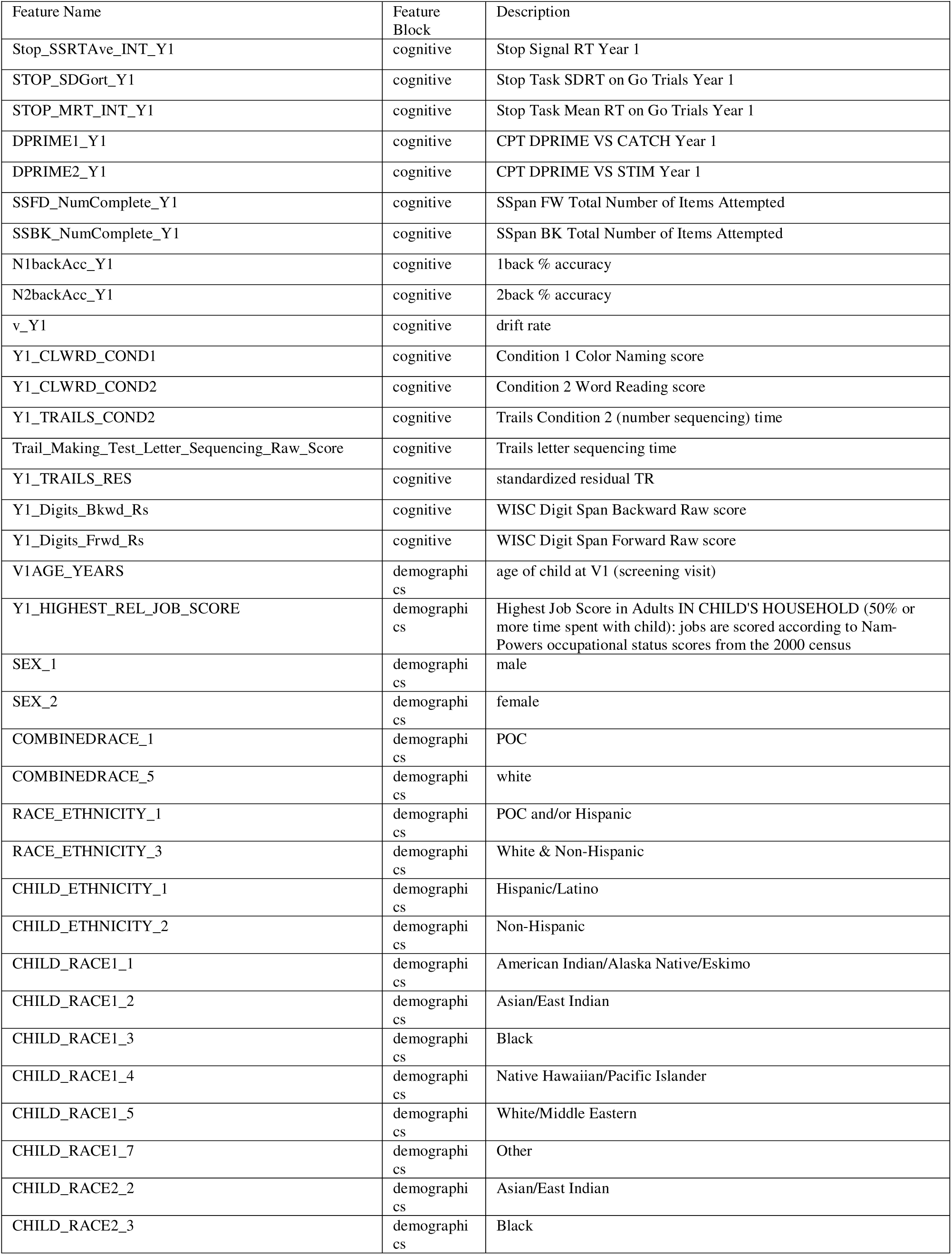
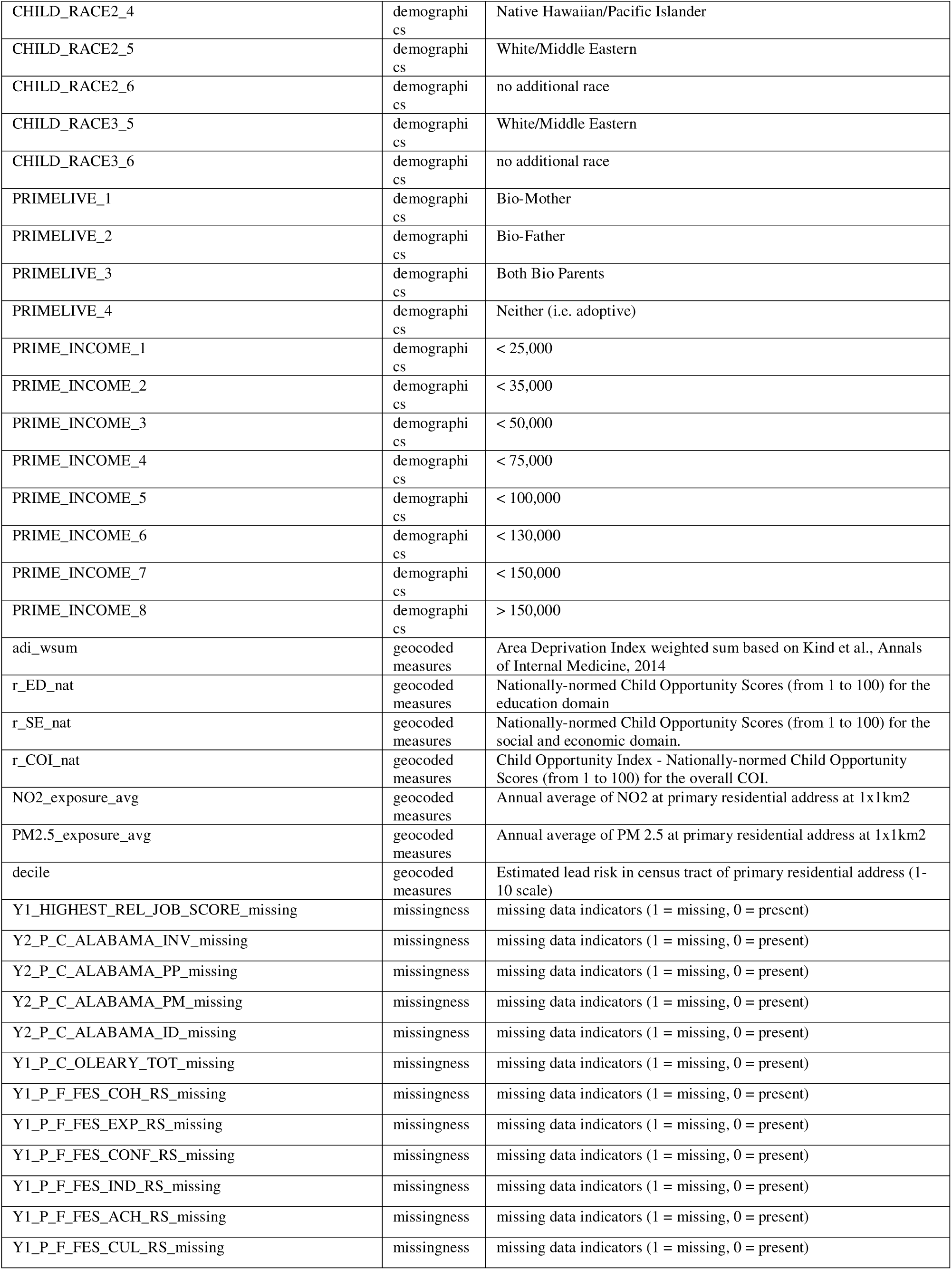
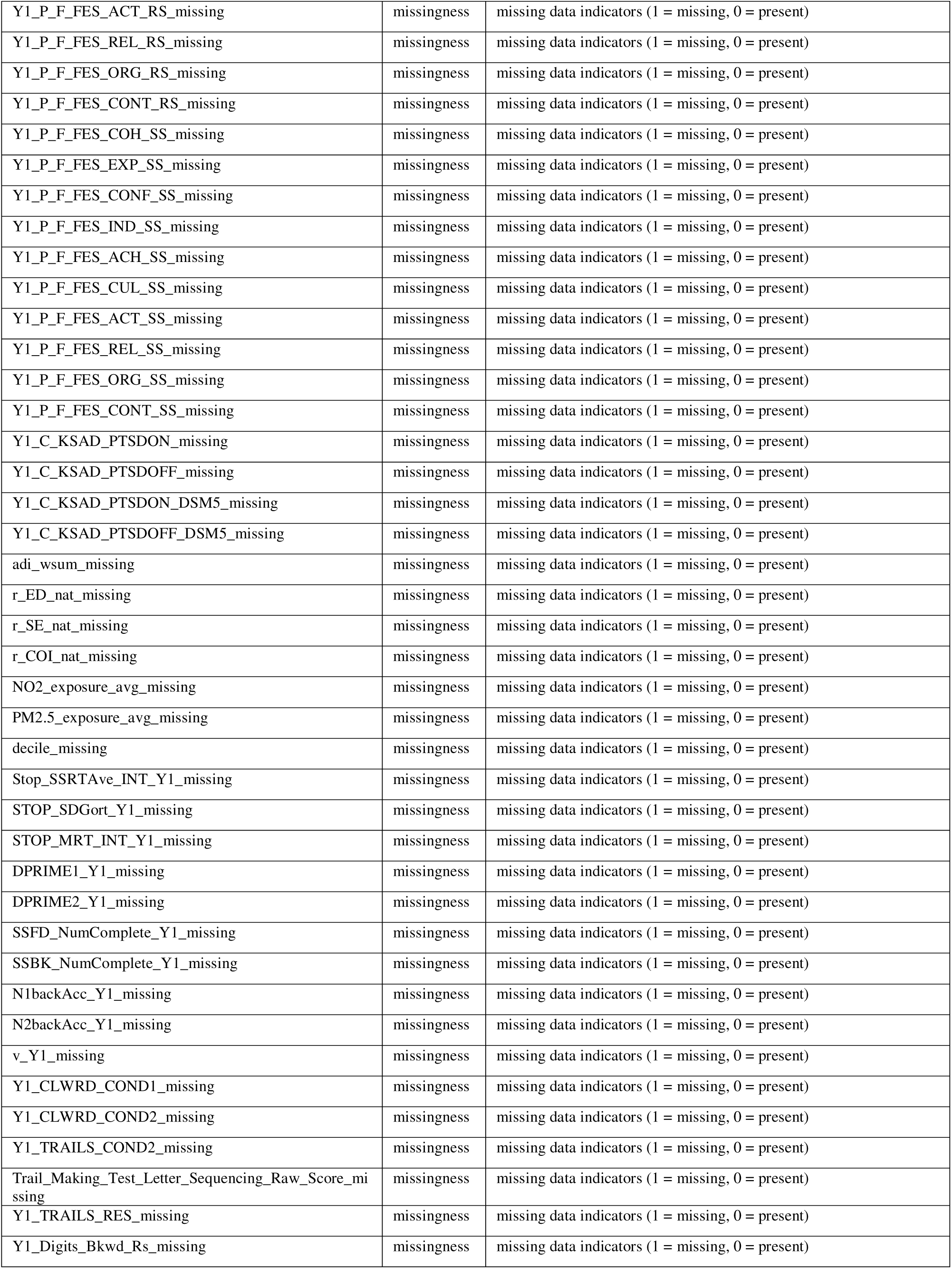
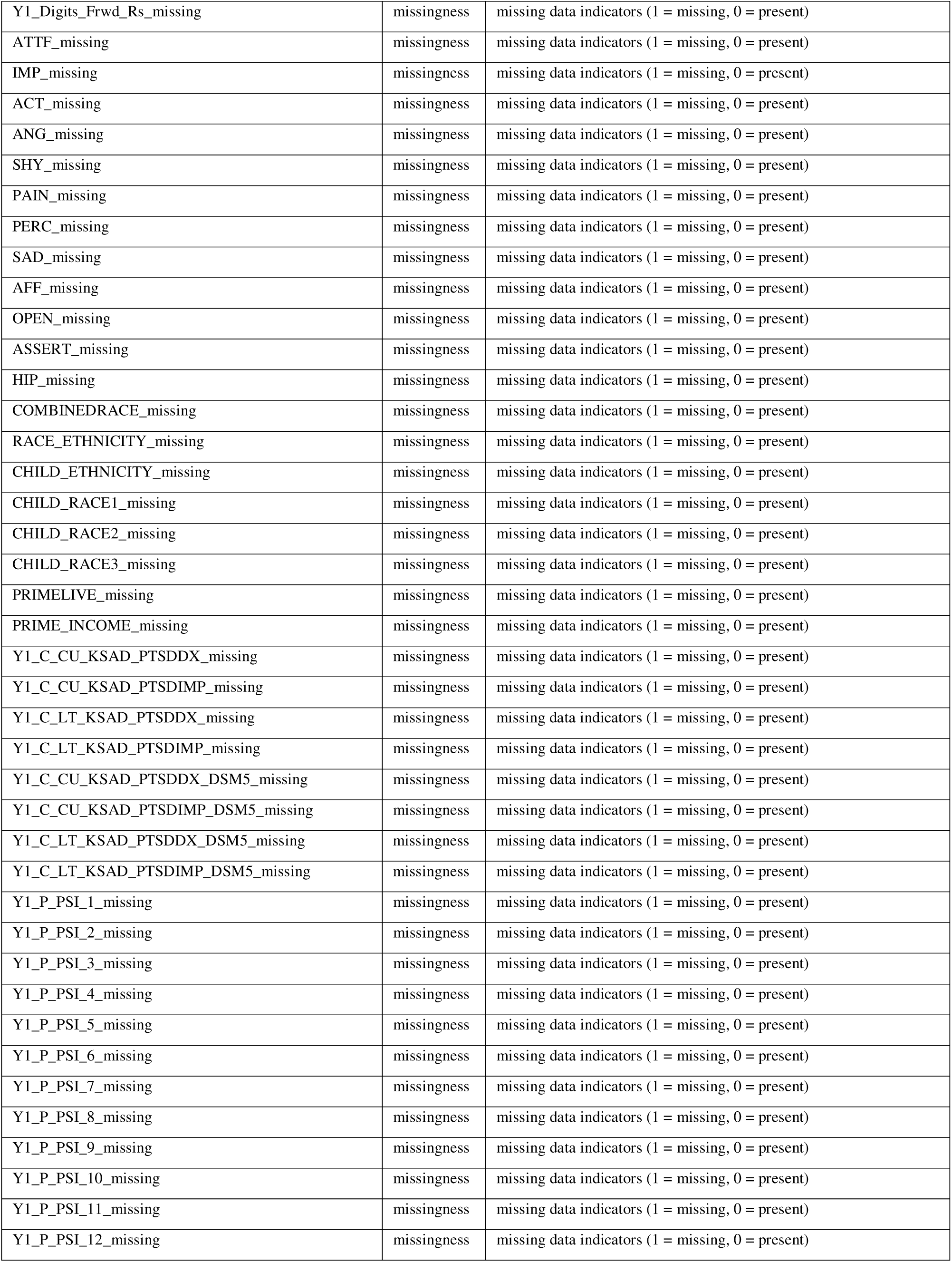
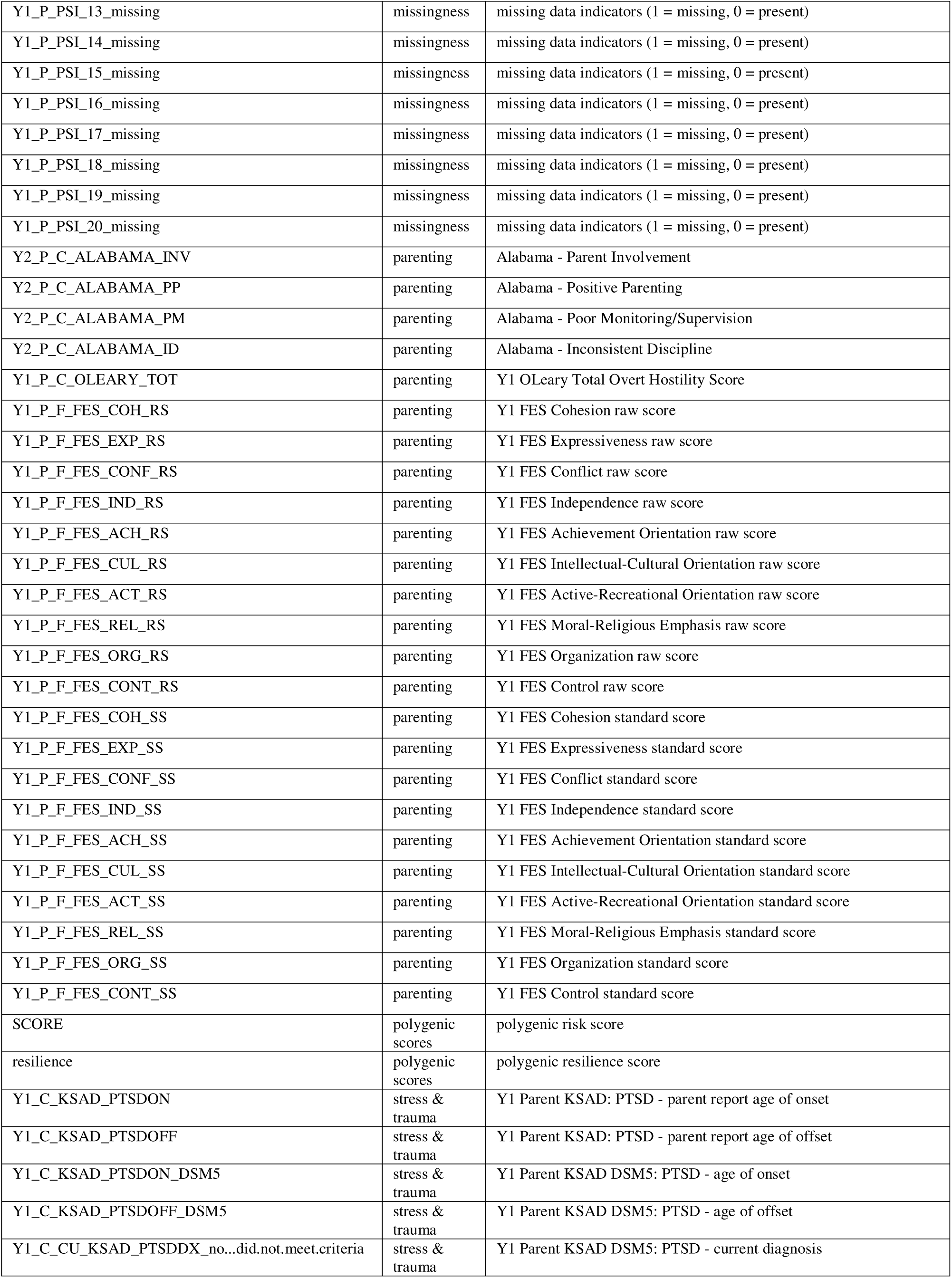
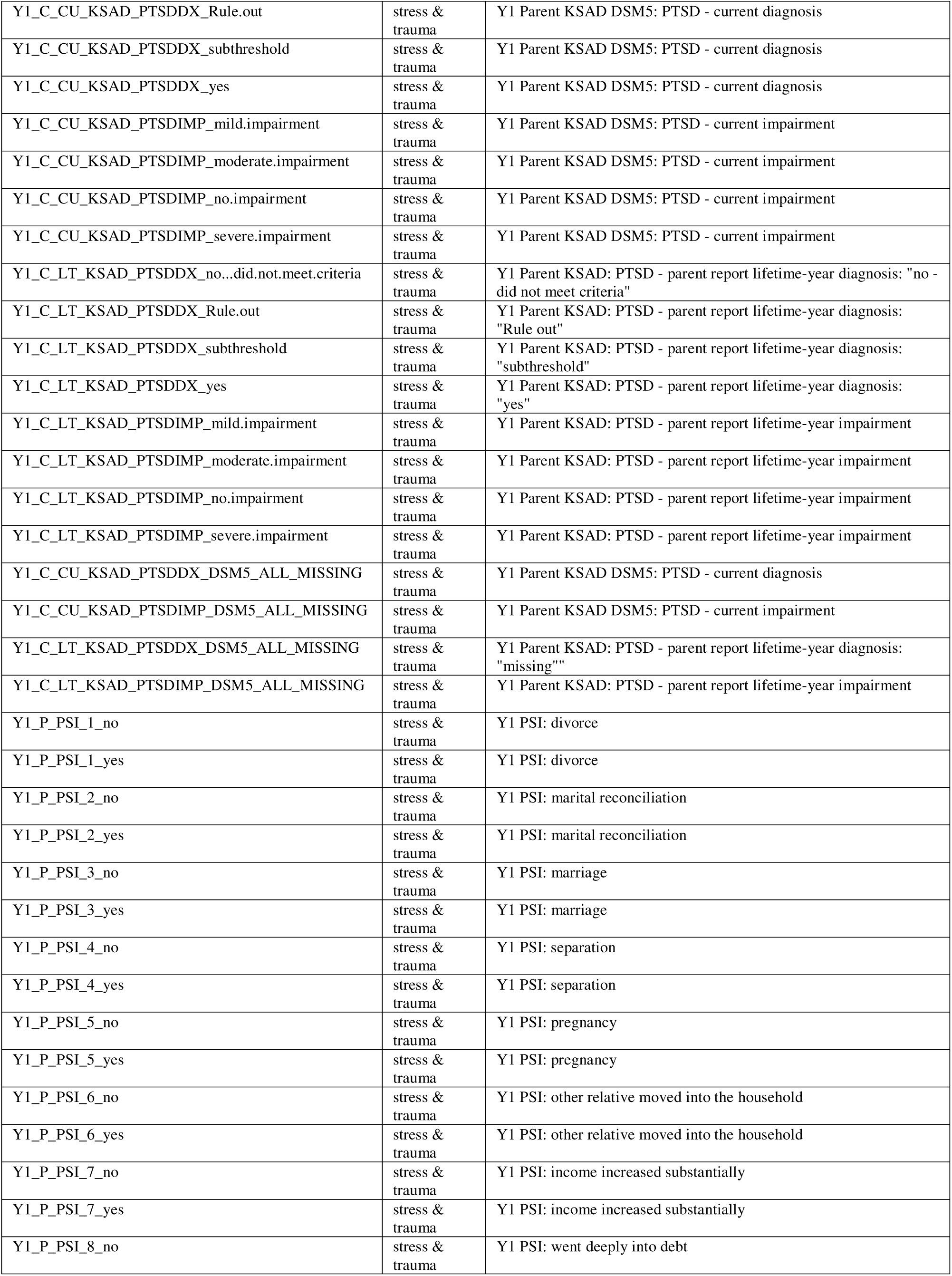
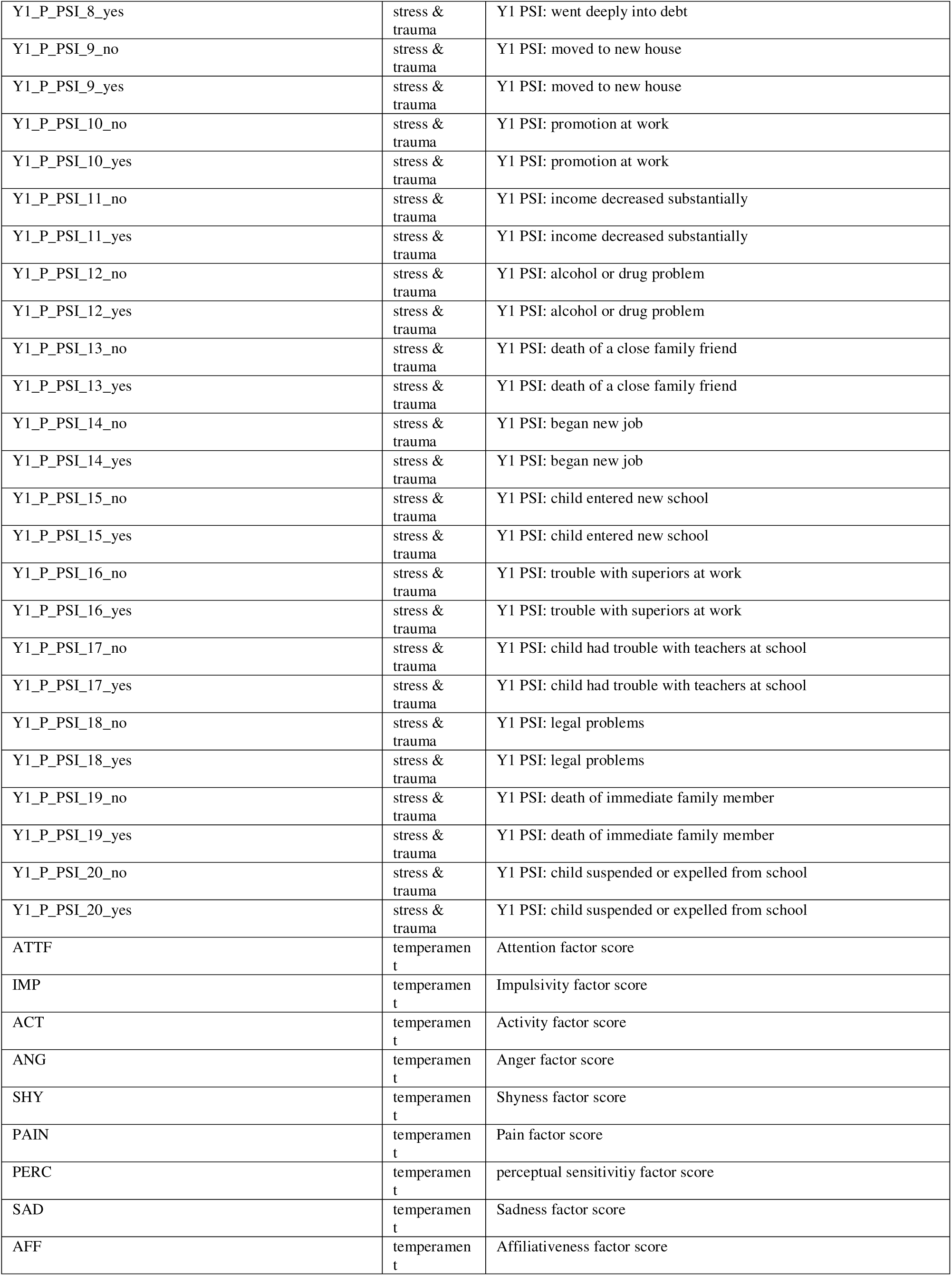
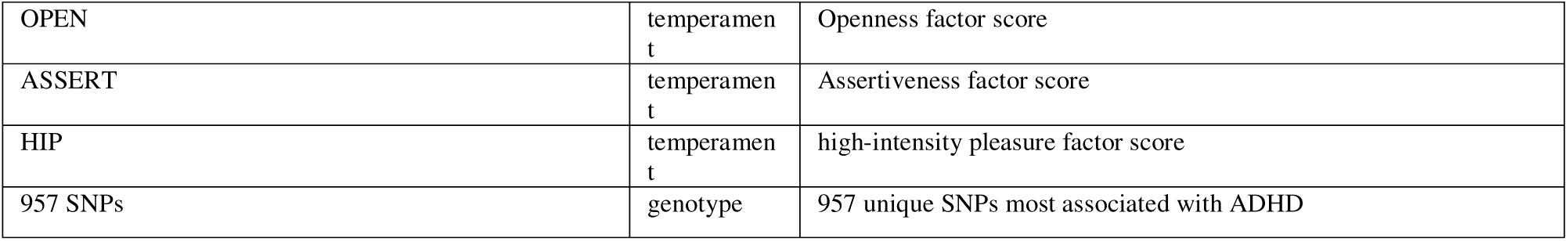
Feature list, block inclusions, and feature descriptions.

**Supplementary Table 2:** Hyperparameter Ranges.

| Supplementary Table 2: Hyperparameter Ranges |  |
| --- | --- |
| Hyperparameter | Range |
| Epochs | 5 - 50 |
| Include genomics | True/False |
| If genomics == True, number of correlated gwas rows | 1 - 77 |
| If genomics == True, number of gene set rows | 1 - 100 |
| If genomics == True, p threshold for SNP inclusion | 0.0001 - 0.01 |
| If genomics == True, convolutional blocks | 1 - 3 |
| If genomics == True, convolutional filters | 10 - 100 |
| If genomics == True, filter width | 1 - 20 |
| If genomics == True, pool width | 1 - max pool width (based on input dimension and number of blocks) |
| Include polygenic scores | True/False |
| Include demographics | True/False |
| Include parenting | True/False |
| Include stress & trauma | True/False |
| Include geocoded measures | True/False |
| Include cognitive | True/False |
| Include temperament | True/False |
| Include missingness | True/False |
| Dense layers | 1 - 3 |
| Dense layer X nodes (optimized separately for each dense layer) | 5 - 100 |
| Dense layer X dropout (optimized separately for each dense layer) | 0 - 0.5 |
| Learning rate | 1e-6 - 1e-3 |
| Gradient reversal weight | 0.001 - 2 |

**Supplementary Table 3:**
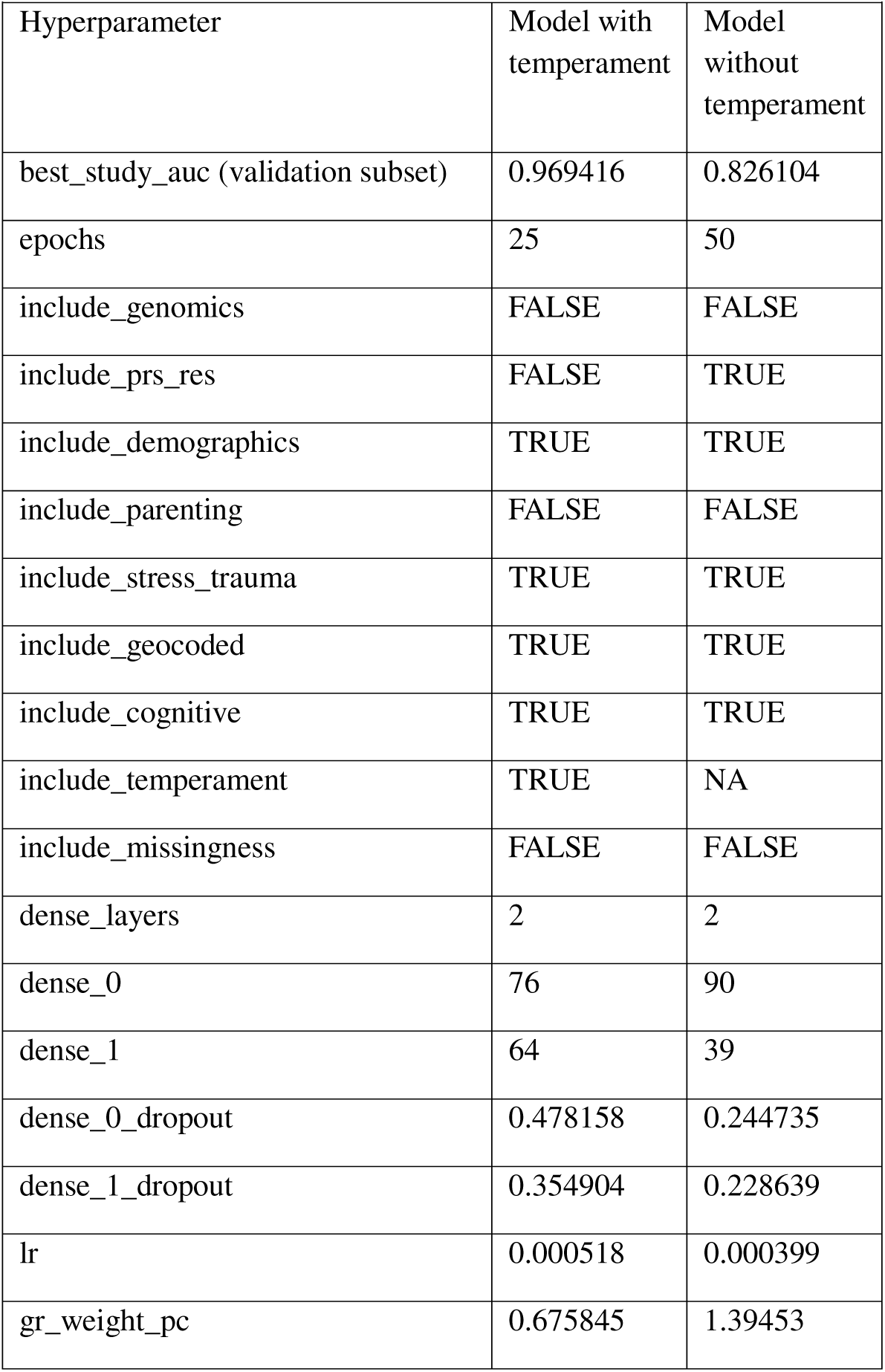
Selected Hyperparameters. See Supplementary Excel file for Supplementary Tables 4-8

| Supplementary Table 3: Selected Hyperparameters |  |  |
| --- | --- | --- |
| Hyperparameter | Model with temperament | Model without temperament |
| best_study_auc (validation subset) | 0.969416 | 0.826104 |
| epochs | 25 | 50 |
| include_genomics | FALSE | FALSE |
| include_prs_res | FALSE | TRUE |
| include_demographics | TRUE | TRUE |
| include_parenting | FALSE | FALSE |
| include_stress_trauma | TRUE | TRUE |
| include_geocoded | TRUE | TRUE |
| include_cognitive | TRUE | TRUE |
| include_temperament | TRUE | NA |
| include_missingness | FALSE | FALSE |
| dense_layers | 2 | 2 |
| dense_0 | 76 | 90 |
| dense_1 | 64 | 39 |
| dense_0_dropout | 0.478158 | 0.244735 |
| dense_1_dropout | 0.354904 | 0.228639 |
| lr | 0.000518 | 0.000399 |
| gr_weight_pc | 0.675845 | 1.39453 |

**Supplementary Table 9.**
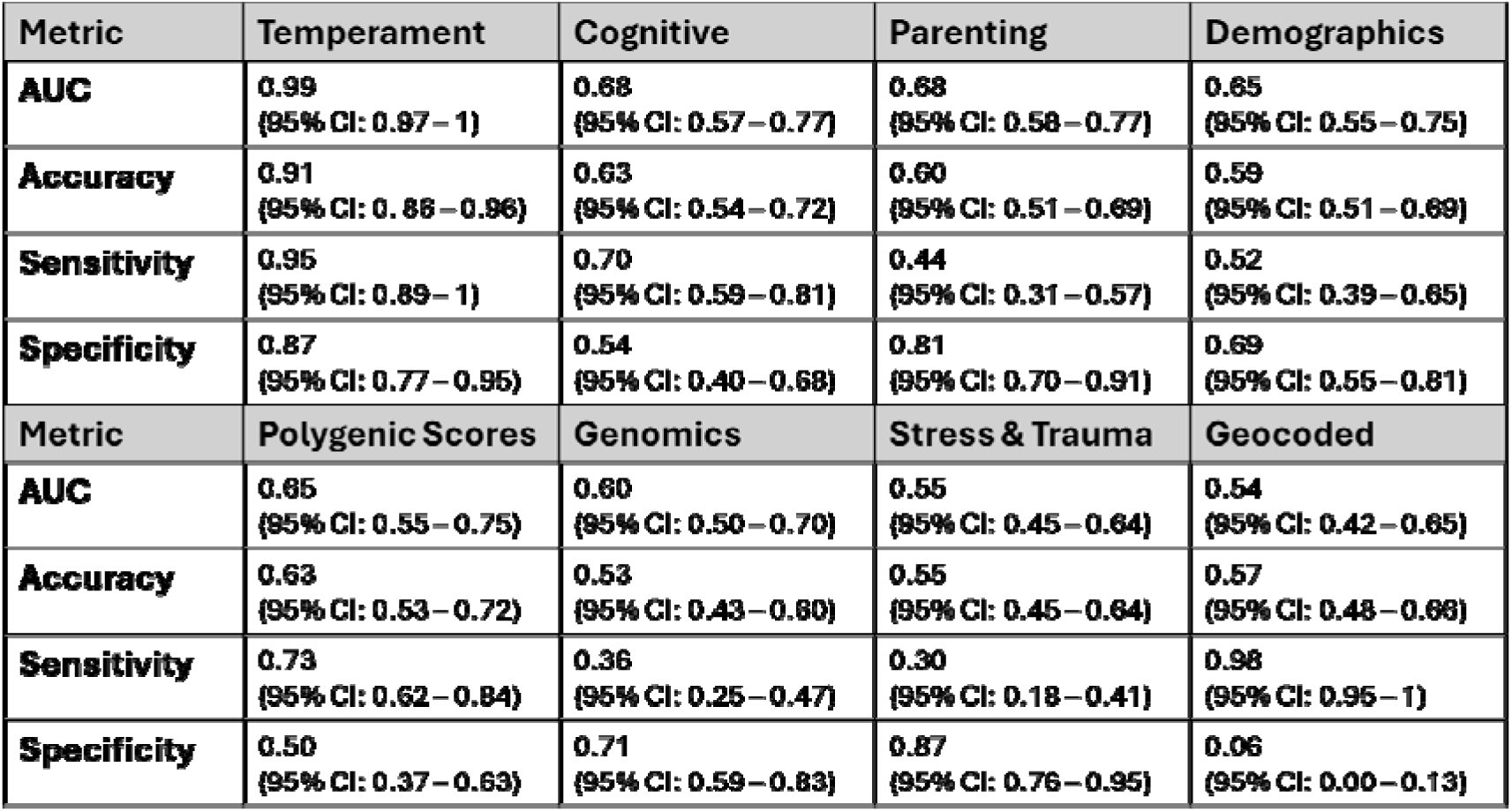
Individual-block model performance classifying diagnostic team ADHD diagnosis.

**Supplementary Figure 1.**
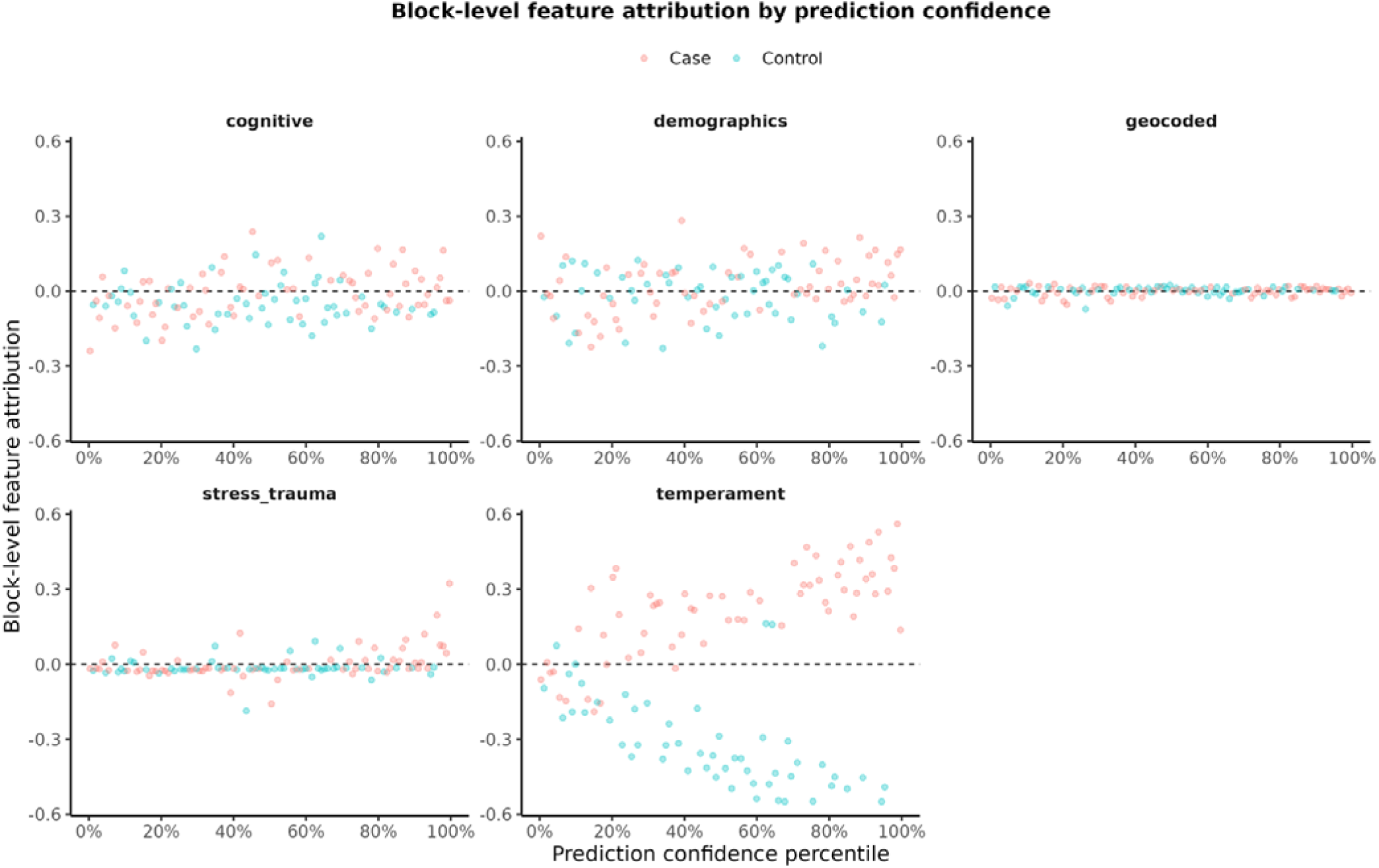
Block-level feature attribution by prediction confidence. Scatter plots showing directional block-level feature attributions across prediction confidence percentiles for each feature block in the temperament inclusive model. Block-level attributions were calculated as the directional proportion of total attribution within each individual, with positive values indicating evidence predicting ADHD risk and negative values indicating evidence predicting less ADHD risk. Temperament showed the clearest separation by diagnostic status, with increasingly positive attribution among cases and increasingly negative attributions among controls at higher prediction confidence.

**Supplementary Figure 2.**
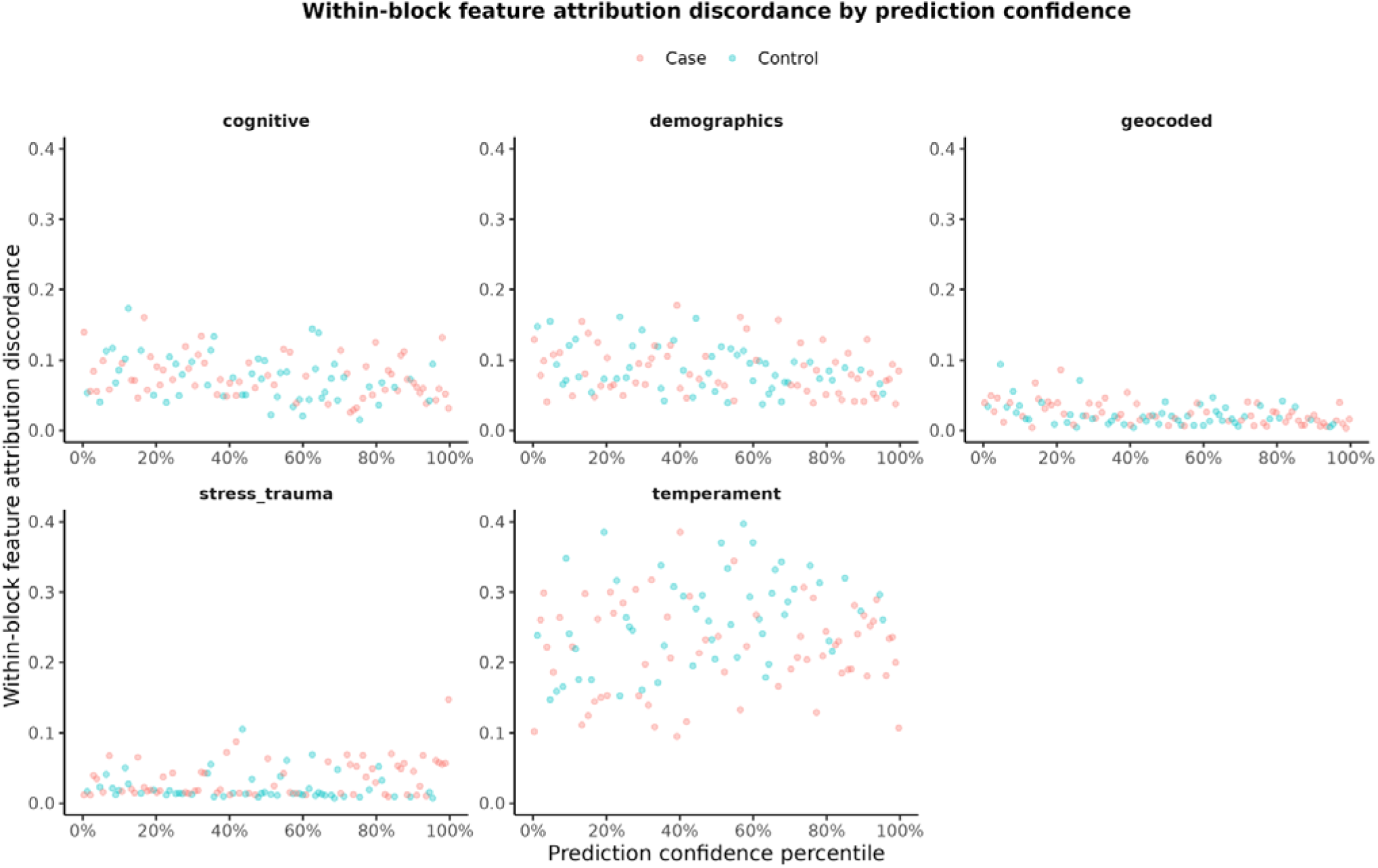
Within-block feature attribution discordance by prediction confidence. Scatter plots showing the relationship between prediction confidence percentile and within-block feature attribution discordance in each feature block. Within-block discordance was defined as the range between the most case-predictive and most control-predictive feature attribution within each block for each individual, with larger values indicating greater disagreement among features within that block for a given individual.

